# From Body to Brain and Back: Multimodal Evidence for Interoceptive Alterations in Schizophrenia Spectrum Disorders

**DOI:** 10.64898/2026.01.13.26344024

**Authors:** Deniz Yilmaz, Lena Deller, Johanna Spaeth, Nina Gottschewsky, Berkhan Karsli, Genc Hasanaj, Linda Sagstetter, Nina Theis, Miriam Zuliani, Annemarie Weibel, Jasmin Jannan, Julia Segerer, Mona Hussain, Vladislav Yakimov, Joanna Moussiopoulou, Antonin Fourcade, Daniel Keeser, James Kilner, Lukas Roell, Michael Gaebler, Arno Villringer, Andrea Schmitt, Isabel Maurus, Peter Falkai

## Abstract

When the brain and body misalign, emotional experience and sense of reality can be disrupted. Although such atypical experiences are central to schizophrenia spectrum disorders (SSD), interoception, processing of internal bodily signals, remains poorly understood in individuals with SSD, particularly across subjective, behavioural, and neural domains. We tested whether SSD is associated with convergent alterations across interoceptive domains and whether these relate to clinical symptoms in a cross-sectional observational design. Patients with SSD (*n* = 53) and matched healthy controls (HC; *n* = 60) completed an EEG experiment comprising eyes-closed and eyes-open resting-state recordings and a heartbeat counting task (HCT), followed by self-report measures. Interoceptive accuracy was derived from the HCT, while cortical responses to afferent cardiac signals were indexed by heartbeat evoked potentials (HEP). In individuals with SSD, subjective interoceptive awareness was altered, characterised by impaired regulation and negative bodily appraisal, alongside elevated depersonalization. At the behavioral level, interoceptive accuracy was marginally lower. HEP positivity was attenuated, most clearly during HCT and, to a lesser extent, in eyes-open rest, over centro-parietal regions, consistent with context-dependent alterations in cortical processing of afferent bodily signals. Symptom–interoception links were largely modest, with depersonalization emerging as the most consistent correlate of clinical severity, suggesting that interoceptive disturbances in SSD may reflect a trait-like, disorder-central alteration. Together, these findings indicate that SSD involves pervasive disturbances in bodily self-experience, marked by impaired interoceptive regulation and by context-dependent neural attenuation during interoceptive engagement. This profile highlights interoceptive dysfunction as a disorder-central feature with potential prognostic and therapeutic relevance.

## 1. Introduction

Picture this: you are going about your day when you suddenly notice your heart racing. There’s no clear reason or danger, yet an ominous sensation takes over, as if someone is out to harm you. This heightened awareness of internal signals, paired with a tendency to misinterpret them, reflects the positive symptoms of schizophrenia spectrum disorders (SSD), such as delusions or hallucinations (Ardizzi et al., 2016a; Pezzulo, 2014; Sakson-Obada et al., 2018; Sterzer et al., 2018; Weilnhammer et al., 2025). Conversely, imagine a reduced awareness of your body, where seemingly exciting external events do not trigger a perception of bodily arousal. This can lead to social withdrawal, lack of motivation, or an inability to feel pleasure, exemplifying negative symptoms of SSD (Jeganathan et al., 2025; Koreki et al., 2021; Yao & Thakkar, 2022a). These context-incongruent experiences highlight changes in interoception, the processing of internal bodily signals, such as heartbeat and breathing (Allen & Tsakiri, 2019; Khalsa et al., 2018; Yao & Thakkar, 2022a), and their potential role in the symptomatology of SSD.

Interoception forms a core component of bodily self-consciousness (BSC), which is frequently altered in SSD and has been described as disembodiment (Irarrázaval, 2015; Martin et al., 2016; Stanghellini, 2009). Different models have been put forward to explain disembodiment. For instance, for several decades, the comparator model has posited a deficit in SSD in distinguishing between sensations originating from the external world, known as exteroception, and self-generated sensations (Frith, 2012; Synofzik et al., 2008). Several cardinal symptoms including hearing voices (Seal et al., 2004; Swiney & Sousa, 2014), delusions of control (Frith, 2012), and somatic passivity (Orepic et al., 2021), have been explained by this model. Ample research inspired by the comparator model has examined alterations in the perception–action loop in SSD, particularly during interactions with the external world, and linked these disruptions to self-disturbances (Möller et al., 2021; Seth, 2013; Voss et al., 2010). Although the perception of internal bodily sensations, i.e., interoception, also plays a key role in shaping BSC (Perry et al., 2019), its alteration and contribution to disembodiment and the broader symptomatology remains comparatively underexplored.

Interoception can be empirically measured on multiple levels (Suksasilp & Garfinkel, 2022), among them: self-reported interoception (i.e., the subjective awareness of interoceptive signals), interoceptive accuracy (i.e., the objective ability to perceive interoceptive signals), and cortical processing of interoceptive signals. The latter can be operationalized using electroencephalography (EEG) by measuring the cortical evoked response time-locked to cardiac activity, termed heartbeat evoked potentials, HEP (Al et al., 2020; Fourcade et al., 2024; Marshall et al., 2022; Schandry et al., 1986). HEP amplitude has been interpreted as reflecting attentional allocation to interoceptive bodily signals relative to exteroceptive input (Petzschner et al., 2018).

Multilevel interoceptive alterations have been reported across a range of psychiatric conditions, underscoring the relevance of bodily signal processing for emotion and self-related disturbances. Despite the well-documented role of interoceptive deficits as a mental health risk factor (Brewer et al., 2021; Khalsa et al., 2018; Schoeller et al., 2024), there is surprisingly little empirical work addressing this in SSD. Alterations in self-reported interoception in persons with SSD have been reported, yet the direction of these effects varies substantially across studies. Specifically, reports range from elevated interoceptive noticing with reduced sustained attention (Koreki et al., 2021) to globally reduced (Torregrossa et al., 2022) or increased (Damiani et al., 2023) self-reported interoceptive awareness. It remains unclear whether these inconsistencies reflect global shifts in interoceptive awareness or differential alterations across specific facets, such as noticing versus regulation, possibly relating to distinct functional processes. Furthermore, several studies report reduced cardiac interoceptive accuracy in SSD compared to HC at the behavioral level (Ardizzi et al., 2016b; Koreki et al., 2021; Torregrossa et al., 2022), yet its association with symptom severity remains inconclusive. Some studies have found that increased interoceptive accuracy predicts reduced negative symptom severity (Koreki et al., 2021), while others report an association between higher interoceptive accuracy and more severe grandiosity symptoms (Ardizzi et al., 2016b), highlighting its potential clinical relevance and the multifaceted nature of these associations.

Neuroimaging evidence based on EEG remains limited and has yielded heterogeneous findings regarding HEP in SSD. A resting-state study by Koreki et al. (2024) reported increased HEP amplitudes over right frontal regions in SSD compared to HC. The authors interpreted this as an underlying attenuation deficit of afferent autonomic signals relative to top-down predictions, resulting in increased interoceptive prediction errors. In contrast, more recent work in SSD has reported reduced HEP amplitudes and altered interoceptive–exteroceptive modulation during brief attentional tasks (Salamone et al., 2025). Together, these findings raise uncertainty regarding the directionality and contextual dependence of HEP alterations in SSD. Critically, prior studies have either focused on resting-state measures or employed brief attentional manipulations without behavioural verification of interoceptive engagement. By assessing HEP during active interoceptive attention, integrating neural responses with behavioural accuracy and subjective interoceptive awareness within the same sample, and applying strict control for physiological and demographic confounds, the present study provides a framework to clarify the direction and functional interpretation of HEP alterations in SSD and to test whether these effects are context-dependent rather than reflecting a fixed baseline shift.

Against this background, the present study aimed to address the lack of integrated, multilevel investigations of interoception in SSD by jointly assessing subjective interoceptive awareness, behavioural interoceptive accuracy, and neural cardiac processing across contexts within a single, well-characterized sample, with the aim of reproducing and extending prior findings. We hypothesized that, compared to HC, individuals with SSD are characterized by (1.1) altered subjective interoceptive awareness, (1.2) reduced behavioural interoceptive accuracy, and (1.3) aberrant HEP amplitudes during eyes-closed rest, particularly over right-frontal electrodes (Fp2, F4, and F8), based on prior findings by Koreki et al. (2024). Moreover, we expect that (2) interoception measures are associated with global clinical outcomes in patients with SSD. Given the heterogeneous and partly inconclusive prior literature, we formulated hypotheses at the level of global clinical outcomes.

Exploratory analyses examined interoceptive subdomain and symptom-specific effects, testing whether interoceptive alterations across neural, behavioral, and subjective levels converge on shared symptom dimensions or remain dissociable. Finally, we probed neural interoceptive processing during active interoceptive attention while validating interoceptive engagement behaviourally, to explore whether the spatiotemporal expression of HEP alterations in SSD reflects impaired engagement with bodily signals rather than a resting-state abnormality.

## 2. Methods

### 2.1. Participants

#### 2.1.1. Patient Sample

We recruited participants aged 18–65 years with a diagnosis of a schizophrenia spectrum disorder (SSD; ICD-10: F2*; World Health Organization, 1993). A total of 67 SSD participants were recruited, of whom 53 had completed baseline EEG and behavioral data and were included in the present analyses. The SSD sample was drawn from an ongoing longitudinal randomized clinical trial investigating the effects of exercise (BrainTrain; DFG-funded, FA 241/21-1; ClinicalTrials.gov ID: NCT05956327), conducted at the Department of Psychiatry and Psychotherapy, LMU University Hospital, Munich. In the present study, baseline data were analyzed using a cross-sectional observational approach..

All patients provided written informed consent, were on stable antipsychotic medication for at least two weeks prior to study participation, had a total Positive and Negative Syndrome Scale (PANSS) score ≤75, and sufficient German language proficiency. Exclusion criteria included serious suicidal risk, relevant psychiatric, neurological, or medical disorders, current substance abuse, pregnancy or breastfeeding, and contraindications for MRI. Participants received €20 for completing the EEG experiment.

#### 2.1.2. Healthy Control Sample

Sex- and education-matched healthy controls (HC, *n* = 61) were recruited via advertisements in the local community and an institutional email distribution list of the LMU psychiatric hospital. Age matching was additionally targeted; although minor differences remained, both groups had a mean age in their thirties. HC participants received financial compensation (€10–€30, depending on recruitment period).

Exclusion criteria for HC included current or past psychiatric or neurological disorders, use of psychoactive medication, relevant medical illness, pregnancy or breastfeeding, substance abuse, and a Brief Psychiatric Rating Scale (BPRS; Overall & Gorham, 1962) score > 20. Further exclusion criteria are provided in the Supplement.

### 2.2. Design and Procedure

All participants attended a single experimental session for EEG data acquisition. HC first completed the BPRS to confirm eligibility. Simultaneous EEG, ECG, and respiration recordings were then obtained for both groups during resting-state eyes-closed and eyes-open conditions (five minutes each), followed by a 10-minute heartbeat counting task (HCT; Schandry, 1981). During resting-state conditions, participants were instructed to remain relaxed with eyes closed, or fixate on a central cross (eyes-open). During the HCT, participants silently counted their perceived heartbeats with closed eyes without taking their pulse. The experimental session lasted approximately 80 minutes and was conducted during daytime.

Analyses in this paper focused on EEG and ECG data. After the EEG procedure, participants completed questionnaires assessing body perception and interoception. Additionally, patients completed clinical and cognitive assessments within the same week as the EEG session. This design ensured that the EEG and interoception assessments were identical for both groups while capturing broader clinical data in patients (see Figure 1).

**Figure 1.**
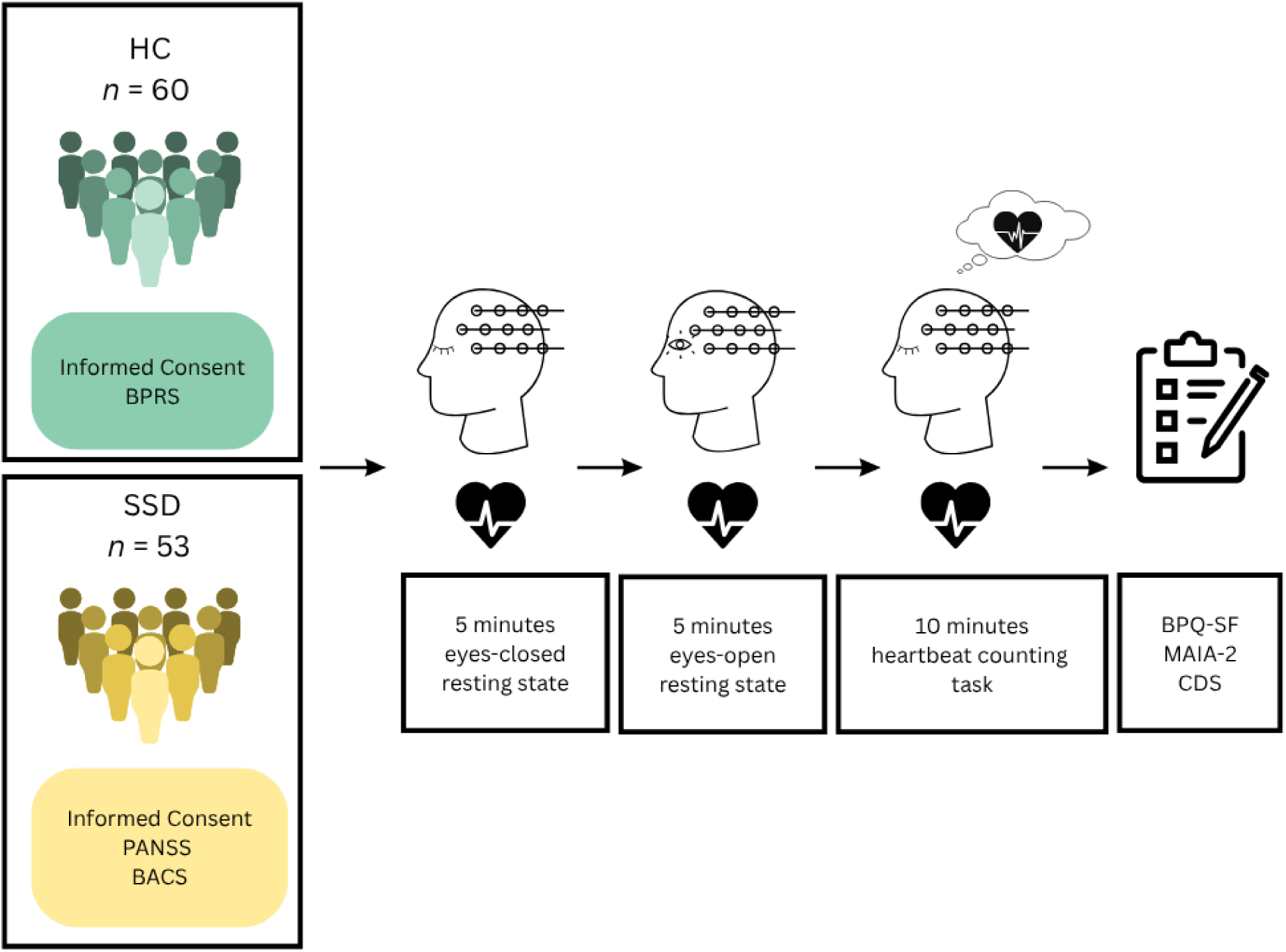
Study Procedure. *Note.* The participants completed the three EEG runs, eyes-closed, eyes-open, and HCT, respectively. Consequently, the questionnaires were administered.

### 2.3. Materials and Measures

Interoception was assessed along three hierarchical dimensions (Suksasilp & Garfinkel, 2022): subjective self-report and beliefs, objective interoceptive accuracy, and neural processing of interoceptive signals (see Supplement).

#### 2.3.1. Self-Report and Beliefs on Bodily Perception

Subjective interoception was assessed using the German-validated versions of the Multidimensional Assessment of Interoceptive Awareness Version 2 (MAIA-2; Eggart et al., 2021) and the Body Perception Questionnaire–Short Form (BPQ-SF; Cabrera et al., 2018). The MAIA-2 assesses multiple facets of interoceptive awareness on a 6-point Likert scale, while the BPQ-SF assesses perceived bodily sensations and autonomic symptoms on a 5-point Likert scale, with higher scores indicating greater awareness and symptom frequency.

Additionally, the Cambridge Depersonalization Scale (CDS; Sierra & Berrios, 2000) was administered to assess depersonalization experiences. The CDS evaluates the frequency and duration of derealization, body distortion, and depersonalization experiences. Higher scores indicate greater depersonalization symptoms. All questionnaires assessed experiences over the past month.

#### 2.3.2. Interoceptive Accuracy

Objective interoceptive accuracy was assessed using the Heartbeat Counting Task (HCT; Schandry, 1981; Koreki et al., 2021). HCT comprised nine trials with durations of 25, 35, and 45 s, during which participants silently counted their perceived heartbeats without taking their pulse. Interoceptive accuracy was computed by comparing reported and actual heartbeats across trials using the standard HCT formula:

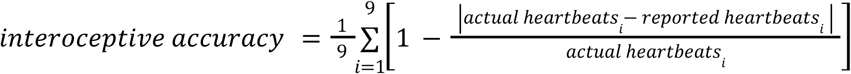

#### 2.3.3. Neural Representation of Interoception

EEG and ECG were recorded simultaneously. HEP were derived as neural markers of cardiac interoception. Mean HEP amplitudes were computed for each electrode and task condition per participant.

#### 2.3.4. Clinical Measures

For the patient sample, symptom severity was assessed using the PANSS (Kay et al., 1987), including the positive, negative, general psychopathology, and total scores. Chlorpromazine-equivalent antipsychotic medication doses (CPZ) were calculated for each patient and included as a covariate in relevant analyses (Leucht et al., 2016). Body mass index (BMI) was assessed and included as a covariate in relevant analyses.

### 2.4. EEG and ECG Data Acquisition and Processing

#### 2.4.1. EEG and ECG Data Acquisition

EEG data was recorded on BrainVision Recorder in a quiet, sound- and electrically-shielded Faraday cage to minimize external noise. We used a 32-channel ActiCAP Ag/AgCl active electrode system and a BrainAmp amplifier (Brain Products GmbH, Germany). The Oz electrode served as a drop-down ECG electrode placed on the left lower back, leaving 31 channels for scalp EEG recording. Signals were sampled at 500 Hz using electrodes arranged according to the International 10–20 system, with the FCz as reference and AFz as ground. During the recording, the electrode impedances were maintained below 25 kΩ. We instructed the participants to relax, minimize movement, and stay awake throughout all recordings.

#### 2.4.2. EEG Preprocessing

EEG preprocessing and analyses were performed in Python using the MNE toolbox v1.9.0 (Gramfort et al., 2014). Data were cleaned using a standardized preprocessing pipeline including bad segment rejection, downsampling to 250 Hz followed by the steps of the PREP pipeline (Bigdely-Shamlo et al., 2015; Fourcade et al., 2024): bandpass and line-noise filtering (0.3–45 Hz; 50 Hz), automated bad-channel detection and interpolation, robust average referencing, and independent component analysis (ICA; Lee et al., 1999; Winkler et al., 2015) to remove ocular, muscular, and cardiac artifacts. Cardiac-related components were additionally identified using an automated ECG-correlation approach.

HEPs were computed by epoching the continuous EEG data time-locked to R-peaks (−250, +550 ms), followed by baseline correction (−125, −25 ms), automated rejection of artifactual epochs, and double R-peak cleaning. Cleaned epochs were averaged per condition (eyes-closed, eyes-open, HCT) to obtain condition-specific HEPs per channel. For the confirmatory analyses, by averaging the signal amplitude in the time window from 450 ms to 500 ms post-R-peak for each channel, we derived the mean HEP amplitudes following Koreki et al. (2024). Further technical details on EEG and ECG processing are provided in the Supplement.

### 2.5. Statistical Analysis

An a priori power analysis (G*Power 3.1) indicated that a total sample size of *N* = 109 was required to detect the effect size reported by Koreki et al. (2024; partial η² = 0.069) with 80% power at α = .05. Descriptive statistics were computed for all relevant variables. Group comparisons between SSD and HC were conducted using independent-samples t-tests (or Mann–Whitney U tests when assumptions were violated) and χ² tests for categorical variables (Fisher’s exact tests when appropriate). See Supplement for full technical details.

#### 2.5.1. Confirmatory Analyses

All confirmatory analyses were preregistered following the model structure described by Koreki et al. (2024) and were evaluated at a two-sided significance threshold of α = .05.

##### 2.5.1.1. Group Differences in Self-Report and Beliefs on Bodily Perception

To test our first hypothesis that self-reported bodily perception differs between groups, separate linear models were fitted for each questionnaire outcome with group as the predictor of interest and age, sex, years of education, and BMI as covariates. *P*-values were adjusted for false discovery rate (FDR) across three questionnaire total scores using the Benjamini–Hochberg (BH) procedure.

##### 2.5.1.2. Interoceptive Accuracy

Our second hypothesis regarding group differences in interoceptive accuracy was tested using a multiple linear regression model with group as the primary predictor; age, sex, education, body mass index, heart rate (HR), and the MAIA Not-Worrying subscale (bodily anxiety proxy) as continuous covariates; and smoking status, caffeine intake, knowledge of one’s own HR as categorical binary covariates, following the covariate structure used by Koreki et al. (2024; see Supplement for covariate rationale and sensitivity analyses).

##### 2.5.1.3. Group Differences in Resting-State HEP in Preregistered Right-Frontal ROIs

Confirmatory HEP analyses were restricted to the eyes-closed condition and preregistered regions of interest (ROIs; Fp2, F4, F8). For each ROI, a multiple regression model was fitted with group as the predictor of interest and covariates capturing demographic, cardiovascular, and behavioral confounds, analogous to the interoceptive accuracy model, with the addition of average QT interval, average R-wave amplitude, and HRV. To ensure model parsimony and stability, covariates were summarized using principal components derived from a Factor Analysis of Mixed Data (FAMD; Austin & Steyerberg, 2017; Jolliffe & Cadima, 2016; Supplement). The resulting model per ROI:

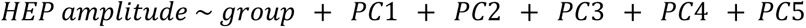

For each ROI, standardized regression coefficients with 95% confidence intervals were extracted to quantify effect sizes adjusted for covariates. FDR was controlled across the three ROIs using the BH procedure. See Supplement for control analyses.

##### 2.5.1.4. Associations With Global Symptom Severity

Partial correlation analyses were conducted within the SSD group to assess associations between symptom severity (PANSS total) and interoceptive measures (questionnaires, interoceptive accuracy, HEP). Correlations were adjusted for age, sex, years of education, BMI, and CPZ. For interoceptive accuracy and HEP correlations, smoking status and caffeine intake were additionally controlled. To account for shared variance among covariates, a FAMD-based dimensionality reduction approach was applied. BH procedure controlled FDR across questionnaires and HEP ROIs.

#### 2.5.2. Exploratory Analyses

##### 2.5.2.1. Questionnaire Subscales

Exploratory group differences in BPQ and MAIA subscales were tested using the same linear model framework as in the confirmatory analyses.

##### 2.5.2.2. Spatiotemporal HEP Differences

To explore group differences in the spatial and temporal distribution of HEPs beyond the predefined ROIs and time windows, we conducted spatio-temporal cluster-based permutation tests comparing SSD and HC separately for the three EEG conditions (eyes-closed, eyes-open, HCT). Analyses were performed across all EEG channels in the 0.25–0.55 s post–R-peak time window, which has been consistently implicated in HEP generation (Coll et al., 2021). Family-wise error rate was controlled at α = .05 using a permutation-based clustering approach (see Supplement).

##### 2.5.2.3. Multivariate Brain–Body–Symptom Associations

Within the SSD group, exploratory Partial Least Squares (PLS) regression was used to examine multivariate associations between interoceptive and physiological measures (X block) and clinical symptom dimensions (Y block: PANSS positive, negative, and general). The X block comprised neural (HEP), cardiac (HR, HRV), behavioral (interoceptive accuracy), and self-report interoceptive measures. PLS was chosen because it does not assume strong intra-block correlations, but instead derives latent components as weighted linear combinations of X variables that maximize covariance with the Y block (Krishnan et al., 2011). To control for demographic and medication-related confounds, all predictors were residualized for age, sex, BMI, years of education, and CPZ. Two latent components were extracted. The contribution of each X variable to the latent components predicting the Y block was quantified using PLS regression coefficients, with stability assessed via jackknife resampling and component significance evaluated using permutation testing (see Supplement).

## 3. Results

Of the 61 HC participants initially recruited, one was excluded from all analyses after post-hoc disclosure of a diagnosed mental health disorder, resulting in a final HC pool of 60 participants. Questionnaire data from one SSD participant were excluded due to incomplete responses. Interoceptive accuracy data were unavailable for two SSD participants (see Supplement for further quality control procedures). See Table 1 and Table S1 for sample characteristics.

**Table 1.** Participant Characteristics.

| Variable | SSD (%) | HC (%) | $\chi^2$ | *p* |
| --- | --- | --- | --- | --- |
| *Demographic and Lifestyle Characteristics (Categorical)* |  |  |  |  |
| Sex (female) | 47.2 | 50 | 0.01 | 0.91 |
| Smoker (yes) | 35.8 | 18.3 | 3.57 | 0.06 |
| Caffeine Intake (yes) | 41.5 | 30 | 1.17 | 0.28 |
| Knows Heartrate (yes) | 14 | 18 | 0.05 | 0.83 |
| Variable | SSD (*M ± SD*) | HC (*M ± SD*) | *t* | *p* |
| *Demographic, Lifestyle, and Clinical Characteristics (Continuous)* |  |  |  |  |
| Age | 38.58 ± 11.77 | 33.72 ± 12.82 | -2.1 | **0.04** |
| Body Mass Index | 29.20 ± 4.44 | 23.29 ± 3.29 | -7.95 | **< 0.0001** |
| Years of Education | 17.47 ± 4.06 | 18.38 ± 3.54 | 1.26 | 0.21 |
| PANSS Total | 48.62 ± 12.37 | - | - | - |
| PANSS Positive | 10.55 ± 3.78 | - | - | - |
| PANSS Negative | 13.68 ± 5.48 | - | - | - |
| PANSS General | 24.40 ± 6.13 | - | - | - |

*Note.* Values are presented as percentages for categorical variables and as mean ± standard deviation for continuous variables.

### 3.1. Confirmatory Results

#### 3.1.1. Group Differences in Self-Report and Beliefs on Bodily Perception

To examine group differences in self-reported bodily perception, we fitted linear models and found that individuals with SSD reported a reduced conscious interoceptive body awareness as measured by MAIA total (β = −0.83, 95% CI [−1.29, −0.36], *p*_adjusted_ = 0.001), alongside increased depersonalization (CDS; β = 0.88, 95% CI [0.41, 1.35], *p*_adjusted_ = 0.001). In contrast, no group differences were observed for BPQ total scores, indicating comparable perceived bodily and autonomic symptom frequency across groups (see Figure 2A and Supplement).

**Figure 2.**
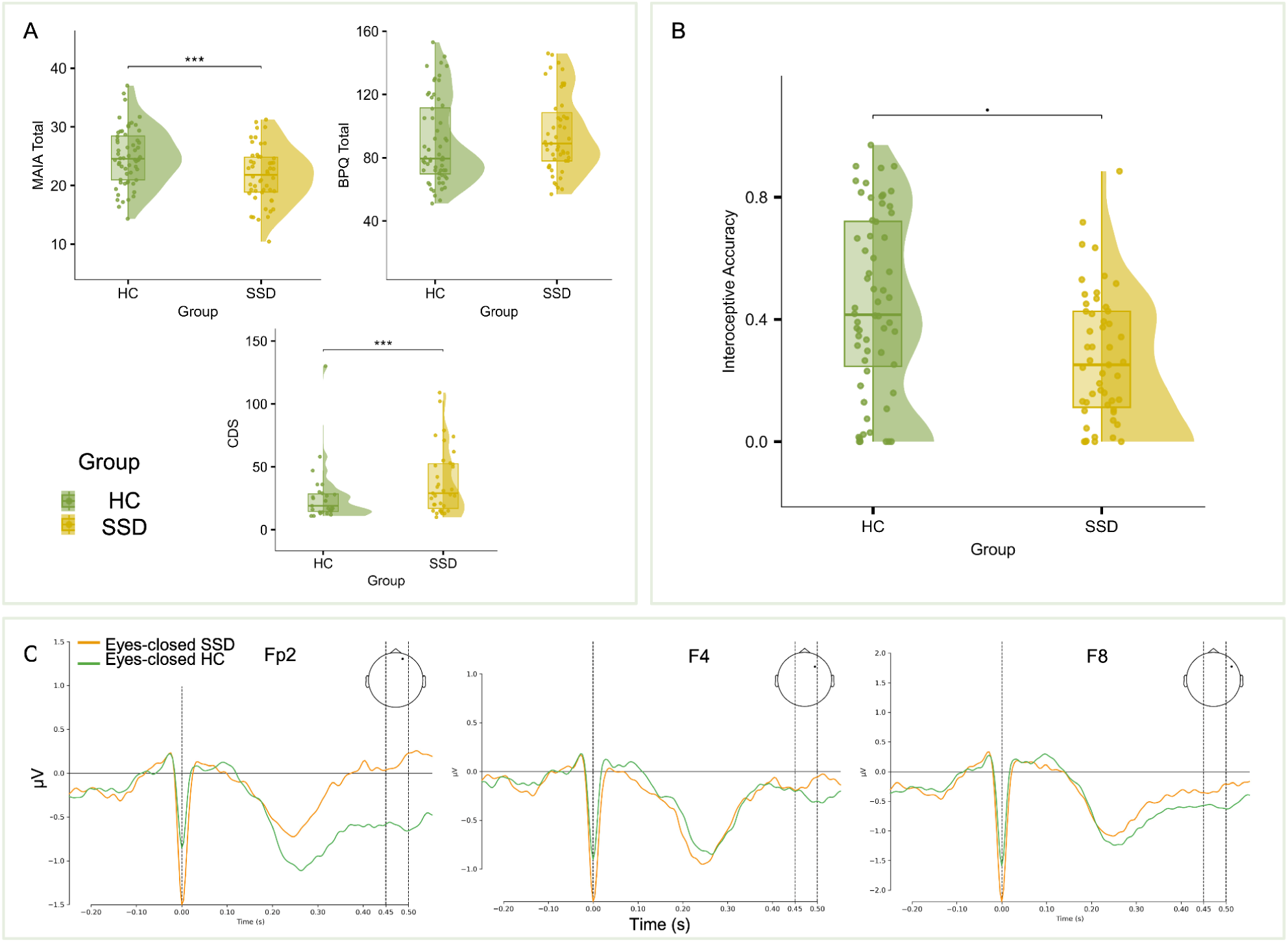
Confirmatory Results: Group Differences in Self-report, Interoceptive Accuracy, and HEP in ROI. *Note.* (A) Group differences for the total scores of MAIA, BPQ, and CDS; pertaining to hypothesis 1.1. (B) Group Differences for interoceptive accuracy; hypothesis 1.2. (C) Mean EEG amplitudes for the ROI (Fp2, F4, F8; respectively) during eyes-closed rest for both groups. The vertical dashed area highlights the time of interest (450-500 ms post-R-peak); hypothesis 1.3. *** : *p* ≤ 0.001; **·** : 0.05 < *p* < 0.10

#### 3.1.2. Group Differences in Interoceptive Accuracy Depend on Model Specification

Individuals with SSD showed a non-significant trend toward lower interoceptive accuracy compared with HC in the preregistered model (β = −0.48, 95% CI [−0.99, 0.02], *p* = 0.058, *R^2^*_adjusted_ = 0.16; see Figure 2B and Supplement for the full model parameters).

In a sensitivity analysis using a more parsimonious model, this group difference reached statistical significance, with participants in the SSD group showing lower interoceptive accuracy compared to HC (β = −0.177, 95% CI [−0.269, −0.084], *t* = −3.77, *p* = 0.0003, *R^2^*_adjusted_ = 0.18). Interoceptive accuracy was positively associated with attentional performance (see Supplement).

#### 3.1.3. No Group Differences in Resting-State HEP in Preregistered Right-Frontal ROIs

No significant group differences in HEP amplitudes were observed after adjustment for covariates at any of the predefined ROI channels: Fp2 (β = 0.35, 95% CI [−0.10, 0.81], *p*_adjusted_ = 0.38), F4 (β = −0.12, 95% CI [−0.57, 0.33], *p*_adjusted_ = 0.60), or F8 (β = 0.13, 95% CI [−0.32, 0.59], *p*_adjusted_ = 0.60; see Figure 2C, Table S5). Therefore, the results did not replicate the results reported by Koreki et al. (2024). There were no group differences in peripheral ECG amplitude, excluding residual cardiac field artifacts (CFA) as a confound (see Supplementary Control Analyses; Figure S1).

#### 3.1.4. Global Symptom Severity Is Selectively Associated With Depersonalization

After covariate adjustments, PANSS total was not significantly correlated with MAIA total (*r* = −0.23, 95% CI [−0.48, 0.06], *p*_adjusted_ = 0.12) or BPQ total (*r* = 0.25, 95% CI [−0.04, 0.44], *p*_adjusted_ = 0.12). However, PANSS total showed a strong positive association with CDS (*r* = 0.55, 95% CI [0.32, 0.72], *p*_adjusted_ < 0.0001; see Figure 3A).

**Figure 3.**
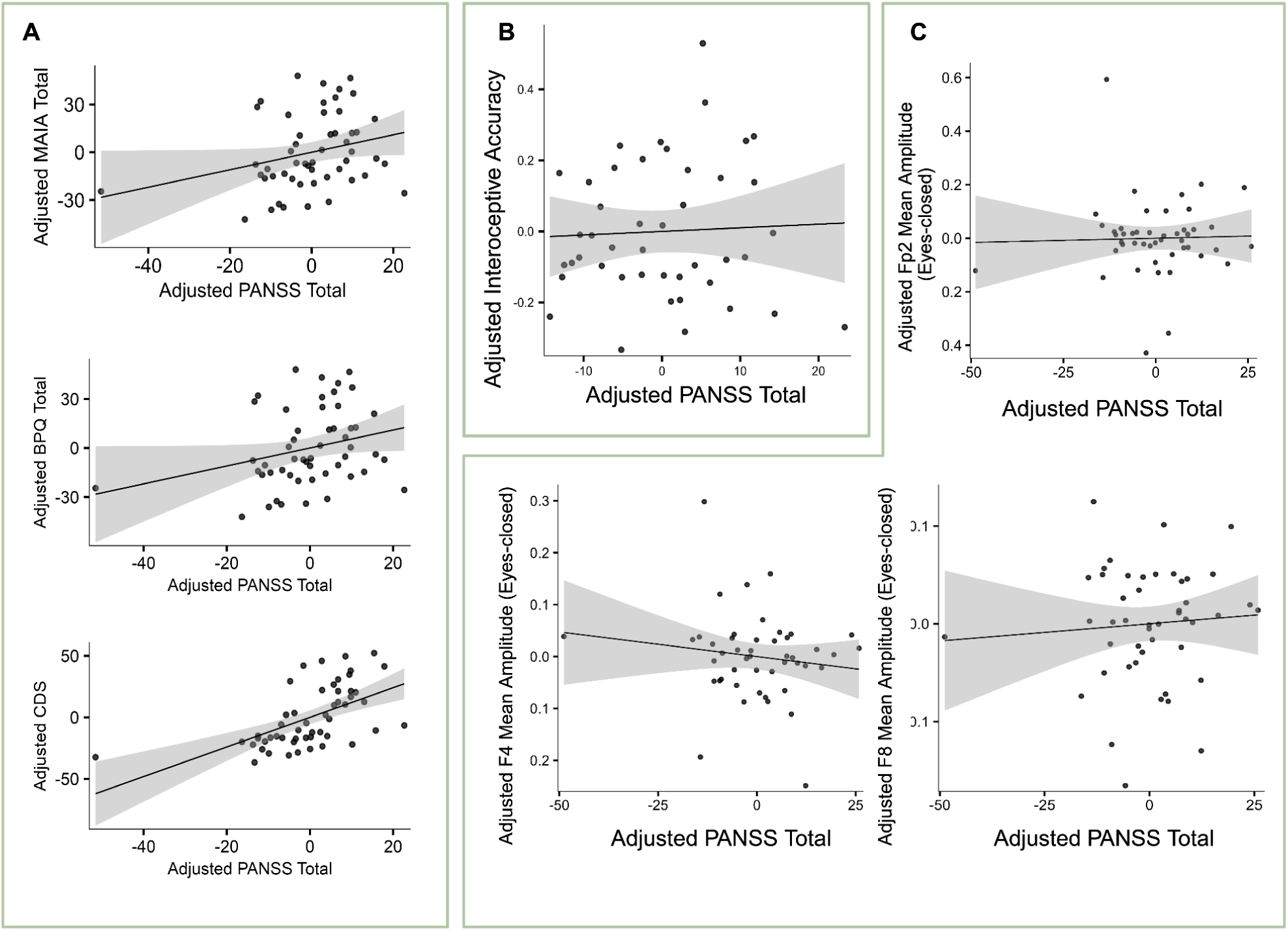
Confirmatory Results: Association between Global Symptom Severity and Interoceptive Domains in SSD. *Note.* Panels show partial Pearson correlations between PANSS total and (A) interoceptive questionnaire totals, (B) interoceptive accuracy, and (C) HEP mean amplitudes at right-frontal ROIs during eyes-closed rest. All analyses were adjusted for age, sex, education, BMI, and CPZ; smoking status and caffeine intake were additionally controlled for interoceptive accuracy and HEP analyses. Shaded bands indicate 95% confidence intervals.

There were no evidence for a relationship between PANSS total and interoceptive accuracy (*r* = 0.11, 95% CI [−0.20, 0.41], *p* = 0.47; see Figure 3B), or HEP amplitude at any ROI: Fp2 (*r* = −0.06, 95% CI [−0.34, 0.23], *p*_adjusted_ = 0.94), F4 (*r* = −0.13, 95% CI [−0.41, 0.30], *p*_adjusted_ = 0.94), or F8 (*r* = 0.01, 95% CI [−0.28, 0.30], *p*_adjusted_ = 0.94; see Figure 3C). See Table S6.

### 3.2. Exploratory Results

#### 3.2.1. Questionnaire Subscales: Selective Alterations in Interoceptive Subdomains

Exploratory analyses of MAIA subscales revealed selective reductions in self-reported interoceptive awareness in SSD participants, particularly in the domains of emotional and regulatory processing (Not Worrying, Attention Regulation, Emotional Awareness, Self-Regulation, and Trusting; all *p*_adjusted_ < 0.05). In contrast, no group differences were observed for the Noticing, Not Distracting, or Body Listening subscales (all *p*_adjusted_ > .35).

On the BPQ, SSD participants showed higher scores on the supra-diaphragmatic reactivity subscale (*p*_adjusted_ = 0.025), indicating increased awareness of upper-body autonomic sensations, whereas no group differences were observed for Body Awareness or Sub-diaphragmatic sensations (see Figure 4A and Supplement).

**Figure 4.**
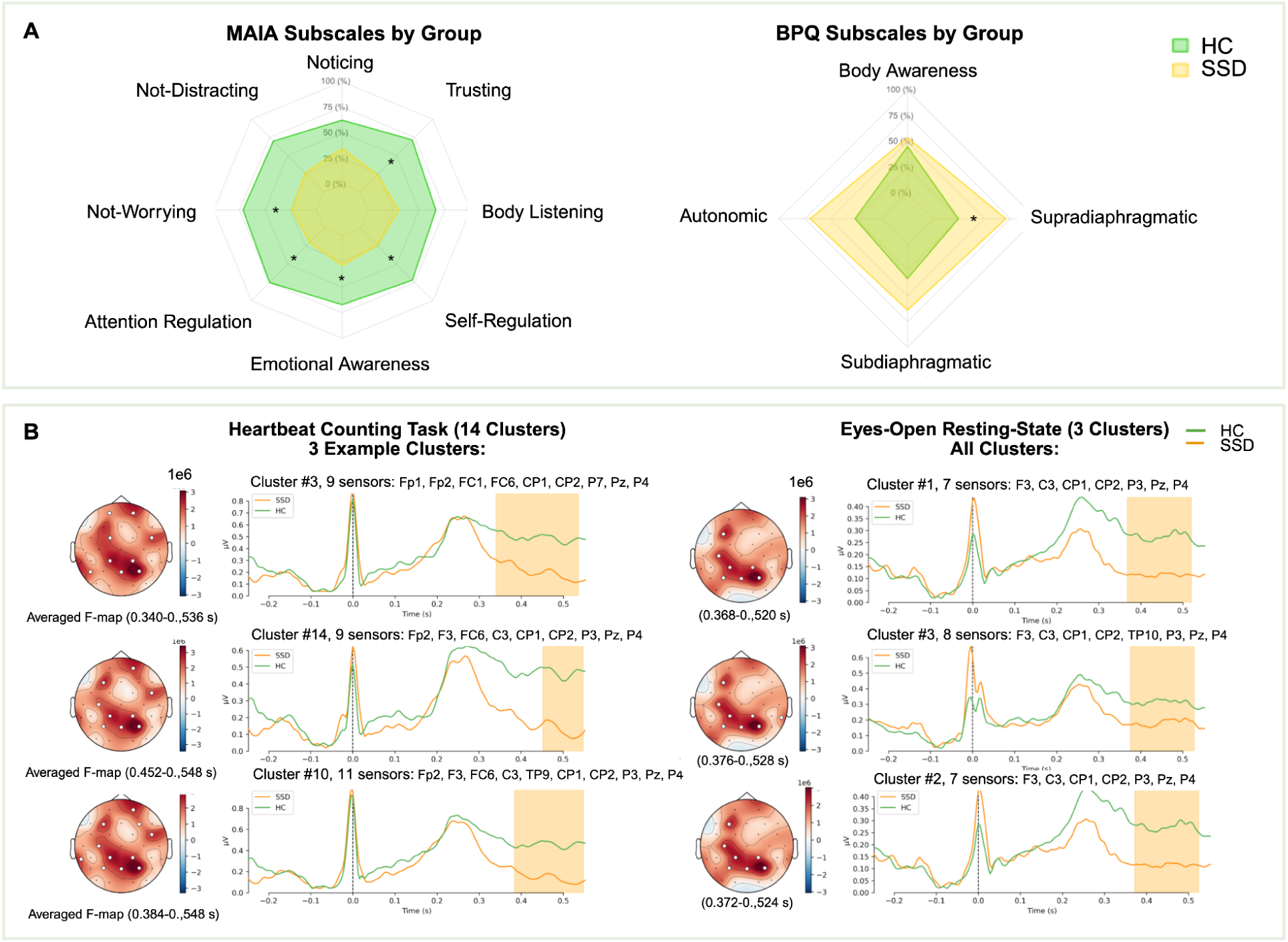
Exploratory Results: Group Differences in Self-Report Subscales and Exploratory Spatiotemporal HEP. *Note*. (A) Group differences in MAIA (left) and BPQ (right) subscales (* : *p* < 0.05). (B) Cluster-based permutation results for the HCT (left) and eyes-open rest (EO; right). During HCT, twelve significant spatiotemporal clusters were identified; for visualization purposes, three representative clusters are shown, as all clusters exhibited highly similar temporal profiles and centro-parietal topographies. EO revealed three significant clusters. Across both conditions, clusters convergently indicated reduced HEP amplitudes in SSD participants, most pronounced approximately 400–500 ms post–R-peak, illustrated using mean Global Field Power (GFP) across sensors contributing to each cluster.

#### 3.2.2. Spatiotemporal Context-Dependent HEP Reductions During Interoceptive Engagement

No significant spatio-temporal HEP clusters were observed in the eyes-closed condition. In contrast, during the HCT condition, 14 significant spatio-temporal clusters revealed reduced HEP positivity in the SSD group, primarily within a centro-parietal topography and spanning approximately 0.30–0.55 s post–R-peak. A similar but less robust pattern was observed in the eyes-open condition, with three significant clusters emerging between 0.37–0.53 s post–R-peak. Thus, group differences in HEP were context-dependent, being most pronounced during interoceptive engagement (HCT), weaker during eyes-open rest, and absent during eyes-closed rest (see Figure 4B, Table S7). See Supplement for exploratory ROI-based group × task analyses.

#### 3.2.3. Multivariate Brain–Body–Symptom Associations

Two latent PLS components were extracted to examine multivariate associations between interoceptive/physiological measures and clinical symptoms. Permutation testing indicated that neither component was statistically significant (Component 1*: p* = 0.74; Component 2: *p* = 0.74), indicating no robust global multivariate brain–body–symptom coupling at the component level.

Exploratory jackknife analyses nevertheless suggested directionally consistent associations, most prominently linking PANSS negative symptoms with higher BPQ supra-diaphragmatic reactivity (*p* = 0.009) and CDS scores (*p* < 0.001), but lower MAIA Trusting (*p* = 0.016). Additional trend-level associations were observed for PANSS positive symptoms (positive trend: interoceptive accuracy, *p* = 0.054; CDS, *p* = 0.086; and MAIA body listening, *p* = 0.055) and general (positive trend: CDS, *p* = 0.054) symptom domains; however, no effects survived corrections (see Supplement, Table S9, Figure S3).

## 4. Discussion

Here, our aims were threefold: (1) to assess cross-sectional interoceptive alterations in SSD across subjective, behavioral, and neural levels, (2) to investigate the clinical relevance of these alterations within the patient sample, and (3) to explore whether interoceptive disturbances show domain-specific, symptom-specific, and context-dependent patterns beyond global effects.

### 4.1. Self-reported Interoception

Our findings support Hypothesis 1.1., demonstrating that self-reported interoception is altered in SSD compared to HC. We showed that SSD scored lower on MAIA, but comparable in BPQ total scores. Although both instruments assess interoception, where MAIA captures regulatory, emotional, and appraisal-related facets beyond mere signal detection, BPQ addresses the frequency of bodily and autonomic symptoms. Concurrently, SSD participants exhibited substantially elevated CDS scores, supporting the prominence of depersonalization and bodily self-disturbances in SSD, aligning with prior work (Humpston et al., 2020; Luque-Luque et al., 2016; Sass, 1987, 2014).

Exploratory subscale analyses indicated that MAIA reductions were facet-specific, driven predominantly by impairments in attentional and emotional regulation of bodily signals. Despite comparable BPQ total scores, SSD participants showed elevated supra-diaphragmatic symptoms, suggesting heightened perception of upper-body autonomic sensations. This pattern aligns with our physiological findings of increased HR and reduced HRV, indicating that heightened peripheral signal awareness may become maladaptive when regulatory and emotional integration processes are compromised.

Together, this profile suggests increased supradiaphragmatic interoceptive salience or hypervigilance (Feola et al., 2024; Howes & Nour, 2016; Navalón et al., 2021; Yao & Thakkar, 2022b), co-occurring with impairments in their appraisal, regulation, and integration into a coherent bodily self, providing a mechanistic link between altered interoception and experiences of depersonalization and disembodiment. This framework helps reconcile previously inconsistent reports on self-reported interoception in SSD by supporting patterns reporting preserved or elevated interoceptive noticing (Damiani et al., 2023; Koreki et al., 2021) alongside impairments in adaptive engagement with bodily signals in SSD (decreased trusting and not-worrying; Salamone et al., 2025; Torregrossa et al., 2022). Such disturbances in interoceptive quality rather than quantity – impaired attention regulation, emotional awareness, and self-regulation in our sample – may contribute directly to bodily self-disturbances characteristic of SSD, including depersonalization. These findings challenge the assumption that greater interoceptive awareness is inherently beneficial and underscore its context-dependent and multidimensional nature (Fan et al., 2025).

Other subdomains, including emotional awareness (Damiani et al., 2023; Salamone et al., 2025) and not-distracting (Koreki et al., 2021; Torregrossa et al., 2022), have shown mixed or contradictory results across studies. Beyond subdomain-specificity, methodological factors likely contribute to heterogeneity in the literature. For example, Koreki et al. used the original MAIA-1 (Mehling et al., 2012), whereas we employed MAIA-2 (Mehling et al., 2018), which includes revised items and improved psychometric sensitivity (Eggart et al., 2021; Scheffers et al., 2024). Alternatively, heterogeneity in interoceptive profiles may reflect distinct subtypes within SSD, an issue requiring larger samples to be resolved.

### 4.2. Interoceptive Accuracy

We found partial support for Hypothesis 1.2. In the preregistered model, group differences in interoceptive accuracy did not reach conventional significance, although the effect was in the expected direction.

Given the relatively high number of predictors specified a priori relative to the available sample size, we additionally evaluated a more parsimonious model to assess the robustness of the group effect under reduced model complexity. This approach was motivated by established concerns regarding variance inflation and reduced estimation stability in overparameterized regression models (Babyak, 2004; Harrell, 2015). In this sensitivity analysis, individuals with SSD showed significantly reduced interoceptive accuracy compared to HC, explaining a greater proportion of variance than the preregistered model. Importantly, none of the additional covariates included in the preregistered analysis significantly predicted interoceptive accuracy, suggesting limited incremental explanatory value beyond group and sex.

Together, these findings indicate directionally consistent evidence for reduced interoceptive accuracy in SSD, converging with prior work using comparable tasks (Jeganathan et al., 2024; Koreki et al., 2021; Torregrossa et al., 2022), with statistical inference sensitive to model complexity in modest samples. Although attentional performance was positively related to interoceptive accuracy, its explanatory power was limited. Furthermore, statistical control for heart rate knowledge and the use of strict sensory-focused task instructions minimizing estimation-related confounds (Desmedt et al., 2018), alongside evidence from less estimation-dependent paradigms (Jeganathan et al., 2024), suggests that reduced interoceptive accuracy in SSD cannot be attributed to domain-general attentional factors alone.

### 4.3. HEP Alterations

Confirmatory analyses (Hypothesis 1.3) revealed no group differences in HEP amplitude during eyes-closed rest at the preregistered right-frontal ROIs (Fp2, F4, F8) after covariate adjustment and FDR correction. Thus, we did not reproduce the resting-state ROI effect reported by Koreki et al. (2024). The absence of group differences in this analysis may partially reflect strict statistical control and adjustment for physiological covariates, potentially attenuating subtle effects. Importantly, however, there were no group differences in ECG amplitude within the HEP time window, reducing the likelihood that residual cardiac field artifacts confounded the EEG findings (see Supplementary Control Analyses).

In contrast to the null findings in the preregistered resting-state ROI analyses, exploratory spatiotemporal analyses revealed context-dependent group differences in HEP expression. During the HCT, SSD participants exhibited markedly reduced HEP positivity, reflected by 14 significant spatiotemporal clusters over centro-parietal regions between 0.40–0.55 s post–R-peak. This topography suggests that group differences extended beyond the preregistered frontal ROIs under sustained interoceptive engagement. A similar but weaker pattern was observed during the eyes-open condition, with three significant clusters within the same time window. This graded pattern – strongest during HCT, weaker during eyes-open rest, and absent during eyes-closed rest – suggests that HEP alterations in SSD depend on task context and interoceptive engagement rather than reflecting a uniform resting-state abnormality.

The absence of group differences during eyes-closed rest likely can be understood in terms of reduced contextual and attentional constraints. Eyes-closed rest represents an unconstrained state in which attention freely fluctuates between bodily sensations and spontaneous, stimulus-independent cognition. In SSD, such states are characterized by altered default-mode dynamics (Northoff & Duncan, 2016) and increased self-generated mental activity (e.g. intrusive inner speech; Alderson-Day et al., 2016), which may divert attentional resources away from bodily signals and reduce the sensitivity of HEPs to detect group-level differences.

In contrast, both eyes-open rest and, more strongly, the HCT impose greater attentional scaffolding and precision demands, either through sustained visual engagement or explicit interoceptive focus. Under these constrained conditions, the interaction between interoceptive and exteroceptive processing becomes more salient, unmasking disrupted interoceptive signal processing in SSD. Additionally, alterations in visual and attentional processing, well-documented in SSD (Boudriot et al., 2024; Levy et al., 2010), may further interfere with the integration of interoceptive and exteroceptive signals under such conditions, amplifying group differences in HEP expression. Together, this pattern suggests that HEP alterations in SSD are not best characterized as a tonic resting-state abnormality, but rather as a context-dependent, dynamically modulated deficit that emerges when interoceptive precision must be actively maintained or integrated with task demands (Ainley et al., 2016; Seth & Friston, 2016).

In line with this interpretation, Salamone et al. (2025) reported lower HEP modulation during interoception relative to exteroception in SSD. We extend these findings by showing that HEP attenuation scales with sustained interoceptive engagement and context across rest and task conditions. The direction of these effects is also consistent with previous work showing attenuated HEP amplitudes in SSD (Koreki et al., 2024). While the authors interpreted their findings as increased HEP in SSD, their plotted waveforms suggest that the effect was characterized by a reduction in negativity, resulting in amplitudes that were closer to zero. Importantly, both their data and ours demonstrated less spatially distinct HEP topographies in SSD (see Figure S4), reflecting a weakened representation of interoceptive signals.

While Koreki et al. (2024) reported attenuated HEPs during eyes-closed rest, their absence here suggests that passive conditions may not consistently reveal this alteration, which instead becomes most apparent when interoceptive precision is behaviorally or contextually engaged. From a predictive coding perspective, this pattern is consistent with altered precision weighting of visceral signals in SSD (Yao & Thakkar, 2022b), such that cardiac inputs are assigned insufficient confidence or precision under increasing inferential demands, resulting in weaker cortical updating of bodily state representations (Barrett & Simmons, 2015; Seth & Friston, 2016; Sterzer et al., 2018).

### 4.4. Clinical Correlations

Confirmatory analyses revealed a robust positive association between depersonalization and total symptom severity. In contrast, none of the other interoceptive measures showed significant associations with PANSS total after covariate adjustment and correction for multiple comparisons. This pattern aligns with prior work reporting weak or inconsistent links between interoceptive markers and global symptom severity in SSD (Jenkinson et al., 2023).

Exploratory multivariate analyses using PLS did not yield significant latent components, indicating the absence of a strong global brain–body–symptom coupling at the component level. Nevertheless, inspection of the exploratory coefficient patterns revealed directionally consistent, symptom-specific associations that may help contextualize the robust depersonalization finding and generate hypotheses for future work, likely reflecting symptom specificity rather than global severity.

The most consistent gradient significantly linked negative symptoms with increased depersonalization, greater supra-diaphragmatic autonomic reactivity, but a decreased trust in bodily sensations. This profile mirrors our self-report findings and suggests an interoceptive–autonomic imbalance characterized by amplified but less reliable bodily signals. Such a pattern may contribute to disembodiment and motivational–affective deficits characteristic of negative symptoms, consistent with accounts of maladaptive regulation based on imprecise interoceptive signals (Ardizzi et al., 2016b; Sedeño et al., 2014; Yao & Thakkar, 2022b). Positive symptoms showed only trend-level associations with greater body listening, higher interoceptive accuracy, and elevated depersonalization. Consistent with prior work (Ardizzi et al., 2016c; Kimhy et al., 2017), this pattern tentatively suggests that in psychosis, compensatory bodily attentional shifts in response to external unreliability may exacerbate bodily detachment when interoceptive signals are imprecise. Nevertheless, these effects require replication and caution is warranted given the lack of correction-surviving effects.

Overall, the weak associations, particularly for neural measures, suggest that many interoceptive abnormalities in SSD may reflect trait-like vulnerabilities, possibly predating illness onset, rather than state markers of symptom severity. Within-patient associations can diverge from group-level deficits: although interoceptive accuracy is reduced on average in SSD, greater accuracy within patients may relate to anxiety or positive symptoms, pointing to partially distinct underlying mechanisms. Importantly, this does not diminish their clinical relevance, as interoceptive processes may still serve as prognostic or treatment-responsive targets (Jenkinson et al., 2023; Yilmaz et al., 2025).

### 4.5. Limitations

Several limitations should be considered when interpreting these findings.

First, the sample size, while powered to detect the preregistered comparisons, remains modest for the multivariate and cluster-based analyses, resulting in a high predictor-to-sample ratio, increasing susceptibility to overfitting and underpowered component tests (Dochtermann & Jenkins, 2011). Accordingly, multivariate findings should be interpreted as exploratory and require replication in larger samples.

Second, the sample comprised post-acute, medicated SSD patients with moderate symptom severity, limiting generalizability to high-risk, acute psychosis, or with treatment-resistant populations. Medication heterogeneity and residual comorbidities may have introduced additional variance not fully captured by CPZ and broad clinical measures.

Third, the cross-sectional design precludes causal inference regarding the origins of interoceptive alterations or the directionality of associations with clinical symptoms. Although key potential confounds were statistically controlled, unmeasured illness-related or contextual factors cannot be fully excluded.

Fourth, interoceptive accuracy was assessed using a single cardiac task. Although we used stringent instructions and verified that heart-rate knowledge did not influence performance, the task remains susceptible to attention- or estimation-related biases (Zamariola et al., 2018; but see Koreki et al., 2021; Minenko et al., 2025). Moreover, both interoceptive accuracy and HEP measures can be influenced by respiratory processes (Zaccaro et al., 2022), which were not explicitly controlled in the present study and should be accounted for in future work. Our findings should be interpreted as reflecting cardiac-specific interoceptive accuracy, particularly given evidence that interoception is not unitary across organ systems (e.g., Banellis et al., 2025; Eimer et al., 2024).

Fifth, the 31-channel EEG montage limited spatial resolution and source localization. ROI- and latency-restricted preregistered analyses may have missed additional effects, while full-scalp cluster findings remain exploratory and require replication with higher-density recordings.

Finally, although task-sequence-related effects (e.g., familiarity, anxiety) cannot be entirely ruled out, the systematic alignment of HEP differences with increasing interoceptive and attentional demands argues against order effects as the primary driver. More generally, the functional interpretation of HEP amplitude remains ambiguous. HEPs reflect cardiac-phase–dependent cortical excitability (Al et al., 2023), but are also influenced by vascular, autonomic, somatosensory, and task-related factors (Jammal Salameh et al., 2024; Luft & Bhattacharya, 2015). Given variability in sources, polarity, and topography across studies, scalp-level effects may obscure region-specific contributions. Accordingly, interpretations in terms of bodily selfhood or interoceptive prediction error remain provisional within the present design.

## 5. Conclusion

Across levels, our findings indicate that interoceptive disturbances in SSD are most robustly expressed at the subjective level. Behavioural and neural alterations were more context-dependent and modest, suggesting that interoceptive dysfunction in SSD is expressed across measurement domains, albeit not uniformly. The robust association between depersonalization and symptom severity underscores disturbances of bodily self-experience as a central clinical correlate of altered interoception in SSD. Together, these results support the view that interoception reflects a disorder-central, yet dynamically modulated, systems-level process underlying bodily self-disturbance in SSD. Interventions targeting interoceptive regulation, such as exercise or other body-based approaches, may therefore hold promise for improving brain–body integration and represent a testable target for future longitudinal and interventional work.

## Data Availability

Explicit permission for data sharing was not included in the informed consent obtained during the data collection phase. As a result, and due to the sensitive nature of the clinical data, we are unable to share the data publicly. However, the data can be made available to individual researchers upon request. Researchers interested in accessing the data may contact us directly to discuss potential arrangements. Other study materials and code are available at https://osf.io/sqa9r/overview and https://github.com/yilmazde/From-Body-to-Brain-and-Back

https://osf.io/sqa9r/overview

https://github.com/yilmazde/From-Body-to-Brain-and-Back

## Acknowledgements

Funding The study was endorsed by the Ministry of Research, Technology and Space (BMFTR) (Bundesministerium für Forschung, Technologie und Raumfahrt) within the initial and the setup phase of the German Center for Mental Health (DZPG)(grant: 01EE2303A, 01EE2303F, 01EE2503F, 01EE2503A to PF). Furthermore, the study was supported by the Else Kröner-Fresenius Foundation with the Research College “Translational Psychiatry” for PF, AS, and IM (Residency/PhD track of the International Max Planck Research School for Translational Psychiatry [IMPRS-TP]), and Max Planck School of Cognition, Max Planck Institute for Human Cognitive and Brain Sciences, Leipzig, Germany, for DY.

