## Supplementary Materials for "From Body to Brain and Back: Multimodal Evidence for Interoceptive Alterations in Schizophrenia Spectrum Disorders"

1. **Supplementary Methods**

**1.1. Participants**

Detailed exclusion criteria for SSD comprised cardiovascular disease, untreated hypertension, infectious disease, cancer, and other major medical conditions. Drug abuse was excluded with the exception of prescribed benzodiazepines. Women of childbearing age were required to use adequate contraception. Additionally, participants were excluded if their residence was too distant from the study center. Patients were recruited via advertisements in local in- and outpatient clinics in Munich, Germany.

Additional exclusion criteria for HC comprised a history of electroconvulsive therapy within the past year, first-degree relatives with psychotic disorders, current cardiovascular disease, and other severe medical conditions. HC participants were recruited via public advertisements and an internal institutional mailing list. The study was in accord with the Declaration of Helsinki and was approved by the ethics committee of the University Hospital, Ludwig-Maximilians-University, Munich.

**1.2. Materials and Measures**

***1.2.1. Self-Report and Beliefs on Bodily Perception***

The MAIA-2 is a 37-item self-report questionnaire that measures multiple facets of conscious interoceptive body awareness. Its subscales assess noticing bodily sensations, the tendency to avoid distraction from sensations, the tendency not to worry about sensations, the ability to regulate and maintain attention to the body, emotional awareness linked to bodily signals, self-regulation through bodily awareness, body listening, and trusting bodily sensations. Each item is rated on a six-point Likert scale from 0 (“never”) to 5 (“always”). These subscales capture multiple facets of interoceptive awareness and bodily self-regulation. While higher scores generally indicate greater awareness or adaptive engagement with bodily sensations, some subscales, particularly Noticing, may reflect a non-linear relationship: very low scores could indicate diminished bodily awareness, whereas very high scores may reflect hypervigilance, both of which could be maladaptive in clinical populations (Bornemann et al., 2015; Mehling et al., 2018).

The BPQ-SF is a 46-item self-report measure that assesses perceived internal bodily sensations and autonomic symptoms. The body awareness domain captures participants’ frequency of perceived bodily signals, while the autonomic symptoms domain is further divided into supradiaphragmatic and subdiaphragmatic reactivity, reflecting responses above and below the diaphragm, respectively. Items are rated on a five-point Likert scale from 1 (“never”) to 5 (“always”), with higher scores indicating greater awareness of bodily sensations and more frequent autonomic symptoms.

***1.2.2. Interoceptive Accuracy***

Objective interoceptive accuracy was assessed using the Heartbeat Counting Task (HCT; Schandry, 1981; Koreki et al., 2021). The HCT comprised three pseudo-randomized runs of three trials with varying durations (25, 35, and 45 seconds) comprising nine trials in total, during which participants silently counted their heartbeats based on internal bodily perception. Accuracy was calculated by comparing counted and actual heartbeats across trials using the following formula, where trials 𝑖 = 1 to 9:

$$interoceptive accuracy =\frac{1}{9}\sum_{i=1}^{9} \left[ 1-\frac{\left| actual heartbeats_{i}- reported heartbeats_{i} \right|}{actual heartbeats_{i}} \right]$$

To ensure the validity of the task, participants were given stringent instructions emphasizing that they should only count heartbeats they could physically feel. They were explicitly instructed not to estimate the number of beats or to rely on external cues such as their pulse, and to report a count of zero if they did not perceive any heartbeats. These instructions were adapted from Desmedt et al. (2020), who recommended modifications to reduce reliance on estimation strategies and enhance the assessment of interoceptive accuracy.

***1.2.3. Clinical Measures***

For the patient sample, symptom severity was assessed using the PANSS (Kay et al., 1987), including the positive, negative, general psychopathology, and total scores. Cognitive performance was screened with the Brief Assessment of Cognition in Schizophrenia (BACS; Keefe et al., 2004); in the present analyses, only the attention subscale was used as a covariate in a sensitivity analysis. Chlorpromazine-equivalent antipsychotic medication doses (CPZ) were calculated for each patient and included as a covariate in relevant analyses (Leucht et al., 2016). Body mass index (BMI) was assessed and included as a covariate in relevant analyses.

### 1.3. EEG and ECG Data Acquisition and Processing

EEG data was recorded on BrainVision Recorder in a quiet, sound- and electrically-shielded Faraday cage to minimize external noise. We used a 32-channel ActiCAP Ag/AgCl active electrode system and a BrainAmp amplifier (Brain Products GmbH, Germany). The Oz electrode served as a drop-down ECG electrode placed on the left lower back, leaving 31 channels for scalp EEG recording. Signals were sampled at 500 Hz using electrodes arranged according to the International 10–20 system, with the FCz as reference and AFz as ground. During the recording, the electrode impedances were maintained below 25 kΩ, as recommended by Brain Products (Emmerling, 2017). We instructed the participants to relax, minimize movement, and stay awake throughout all recordings.

The EEG data were preprocessed and analyzed with Python (v.3.12.8) scripts utilizing the MNE toolbox, version 1.9.0 (Gramfort et al., 2014). During the data collection phase, we made real-time annotations whenever an undesirable event, such as participant verbalizations or technical disturbances, occurred, allowing us to precisely identify and remove these segments during preprocessing. First, we removed these bad segments as well as the breaks during the Heartbeat Counting Task from the recordings. The standard “easycap-M1” montage was applied. We then downsampled continuous data to 250 Hz, followed by adding the R-peak timings as annotations to the data sets. The non-EEG channels were excluded from subsequent EEG-specific preprocessing. Next, we conducted the following steps of the PREP pipeline (Bigdely-Shamlo et al., 2015; Fourcade et al., 2024): bandpass filtering (0.3–45 Hz, zero-phase finite impulse response, FIR, filter), line-noise removal at 50 Hz, detection (find_all_bads() of *pyprep*) and spline interpolation of noisy channels, and robust referencing to average. On average, in HC, the average number of interpolated channels per subject was 8.82 during eyes-closed, 8.94 during eyes-open, and 6.89 during the HCT. In patients with SSD, the corresponding values were 6.45, 8.61, and 4.95 channels, respectively.

Next, we ran an independent component analysis (ICA Extended infomax; Lee et al., 1999) to identify and exclude EEG artifacts due to eye movements, blinks, muscular activity, and cardiac field artifacts (CFA). ICA projection matrices were derived from a duplicate of the original dataset that was subjected to a high-pass filter with a cutoff frequency of 1 Hz, rather than 0.3 Hz, to facilitate the decomposition (Winkler et al., 2015). Rank deficiency correction was implemented by default in the *ICA* class of MNE. The *ICLabel* method automatically labeled the components on the original data preprocessed until ICA. A component was excluded if it was labeled as anything other than “brain” or “other” with a predicted probability above 50%. In addition to iclabel, to reliably eliminate cardiac artifacts in our setting, we used an automated process that identifies ECG-related ICA components based on their similarity to the ECG signal. Specifically, we employed a correlation-based approach, where each independent component was evaluated for its similarity to the ECG signal. The process involved calculating a correlation score for each component and iteratively applying a z-score threshold (threshold='auto', set to 3.0 using the *ica.find_bads_ecg* method on MNE) to identify components that were outliers in terms of their ECG-related activity. This ensures the removal of subtle cardiac artifacts that might not be detected by ICLabel alone. All components identified by ICLabel or as ECG-related were excluded from the original data. This automated method ensured a robust and consistent identification of cardiac artifacts tailored to our dataset. For HC, on average 5.32 (Eyes-closed; heart: 0.75, eye: 2.32, muscle: 2.25), 6.05 (Eyes-open; heart: 0.83, eye: 1.97, muscle: 3.25), and 5.78 (HCT; heart: 0.88, eye: 2.27, muscle: 2.63) ICA components per subject were discarded. For SSD, on average 5.25 (Eyes-closed; heart: 1.16, eye: 2.27, muscle: 1.82), 5.67 (Eyes-open; heart: 1.04, eye: 2.24, muscle: 2.39), and 5.51 (HCT; heart: 1.25, eye: 2.22, muscle: 2.04) ICA components per subject were discarded. The remaining ICA weights were projected back to the data that had been preprocessed up to the ICA stage.

Last, we calculated HEPs. To this end, we epoched the data, starting 250 msec before R-peak and ending 550 msec thereafter. We baseline-corrected the epochs using a baseline period from 125 to 25 ms before the R-peak. Bad epochs were identified and rejected based on a peak-to-peak amplitude threshold of 150 µV for EEG channels. If more than 33% of epochs were rejected, the data was discarded, unless a single channel caused most bad epochs, in which case it was interpolated and epoching were repeated. Furthermore, epochs with more than one R-peak were excluded to ensure that each epoch corresponded to a single cardiac event. For HC, an average of 345.02 (eyes-closed), 345.55 (eyes-open), and 362.53 (HCT) artifact-free epochs per subject were retained for HEP analysis. For patients, an average of 382.48 (eyes-closed), 378.64 (eyes-open), and 395.37 (HCT) artifact-free epochs per subject were retained. The remaining epochs were then averaged to create the evoked object for each condition (eyes-closed, eyes-open, and hct) per channel. For the confirmatory analysis, the time window from 450 ms to 500 ms post-R-peak was marked as the HEP window following Koreki et al. (2024). By averaging the amplitude within this window for each channel, we derived the mean HEP amplitude values per channel.

We used the NeuroKit2 package (v.0.2.10; Makowski et al., 2021) for the ECG preprocessing. First, we ran neurokit2.ecg_process, extracted the continuous ECG signal from the EEG data and downsampled it to 250 Hz. Then, we used neurokit2.ecg_process to automatically detect the R-peaks and the extraction of heart rate (HR), Root Mean Square of Successive Differences (RMSSD) as a measure of heart rate variability (HRV), mean R-peak amplitude, QT interval, and corrected QT interval. Average ECG waveform time-locked to R-peaks was computed per condition using the same baseline correction, epoch range, and time window of interest (450 ms to 500 ms post-R-peak) as the HEP computation. We also extracted the mean and standard deviation of the evoked ECG amplitude within this time window of interest.

We used the NeuroKit2 package (v.0.2.10; Makowski et al., 2021) for the ECG preprocessing. R-peaks were detected automatically, and standard cardiac indices were derived, including HR, root mean square of successive differences (RMSSD) as a measure of HRV, mean R-peak amplitude, QT interval, and corrected QT interval. In addition, R-peak–locked average ECG waveforms were computed per condition, and mean evoked ECG amplitude within the HEP time window was extracted for use in control analyses.

**1.4. Statistical Analysis**

An a priori power analysis was conducted using G*Power 3.1. To achieve 80% power to detect the maximum effect size reported by Koreki et al. (2024; partial η² = 0.069) at α = .05, a total sample size of *N* = 109 was required. Descriptive statistics were calculated for all demographic, physiological, and covariate variables. Group comparisons between SSD and HC were conducted using independent-samples t-tests for continuous variables (Mann–Whitney U tests when assumptions were violated) and chi-square tests for categorical variables (Fisher’s exact tests when expected cell counts were low).

***1.4.1. Covariate Rationale***

Covariates were selected a priori to closely match the analytical approach of Koreki et al. (2024), ensuring methodological comparability and replication fidelity. Specifically, age, sex, years of education, body mass index, heart rate, smoking status, caffeine intake, and knowledge of one’s own heart rate were included to control for demographic, physiological, and behavioural factors known to influence heartbeat perception. The MAIA Not-Worrying subscale was included as a proxy measure of bodily anxiety, consistent with Koreki et al. (2024), and was selected because no standalone anxiety questionnaire was available in the present dataset. Anxiety-related responses to bodily sensations are known to affect interoceptive accuracy and may differ between groups.

***1.4.2. Neural Representation of Interoception***

For the confirmatory HEP analysis, as per our pre-registered protocol replicating Koreki (2024), we only analyzed the eyes-closed condition and built the same models. We ran three multiple regression analyses, one for each of our dependent variables, the pre-defined regions of interest (ROI) channels of Fp2, F4, F8. For this analysis, participants were excluded if more than one of these ROIs had been interpolated, to ensure that the regression analyses captured activity from the target channels rather than being confounded by signals from neighboring electrodes. The models were very similar to the above-described interoceptive accuracy models, having group as the predictor of interest. As covariates, they included age, sex, BMI, years of education, smoking status, caffeine intake, Not-Worrying subscale of MAIA, HR, average QT interval of ECG, average R wave amplitude of ECG, and HRV measured by root mean square of successive differences (RMSSD).

To ensure the statistical power, stability, and parsimony of the primary regression models, we employed a dimensionality reduction technique to synthesize the nine pre-registered covariables. This approach was justified to maintain an appropriate ratio of observations to predictors (109 and 12, respectively, in our case), recommending 15-20 observations per predictor for stable parameter estimation (Austin & Steyerberg, 2017). Given our sample size, a total of 6 predictors were appropriate. Therefore, in addition to the *group* variable, we used the first five resulting principal components (PC) from a Factor Analysis of Mixed Data (FAMD) executed on the eleven covariates. The FAMD was performed using the *FactoMineR* package (Lê et al., 2008), which automatically standardizes the quantitative variables and performs optimal scaling on the qualitative variables. These five PCs cumulatively explained 70% of the variance in the covariates, which is generally considered satisfactory to capture the majority of variance in the dataset (Jolliffe & Cadima, 2016). The resulting models were:

$Fp2 HEP Amplitude \sim group + PC1 + PC2 + PC3 + PC4 +PC5$

$F4 HEP Amplitude \sim group + PC1 + PC2 + PC3 + PC4 +PC5$

$F8 HEP Amplitude \sim group + PC1 + PC2 + PC3 + PC4 +PC5$

For each model, standardized beta coefficients with 95% confidence intervals (CI) were extracted to quantify effect sizes adjusted for covariates. To control the False Discovery Rate (FDR) across the three ROI outcomes, a Benjamini-Hochberg (BH) procedure was applied to the *p*-values of the primary group predictor. Model stability and adherence to assumptions were assessed: inspection of residual distributions (normality and homoscedasticity), leverage and influence diagnostics (Cook’s distance), and variance inflation factors to assess multicollinearity. No substantial violations were observed.

***1.4.3. Correlations***

We conducted partial correlation analyses within the patient sample to assess clinical and interoceptive associations. Partial Pearson correlations, adjusting for age, sex, years of education, BMI, and CPZ, assessed the link between PANSS total and interoceptive parameters (questionnaires, interoceptive accuracy, HEP). For the HEP and interoceptive accuracy analyses, we additionally included smoking status and caffeine intake as covariates. To reduce model complexity and account for correlations among covariates, we performed a FAMD on all covariates, extracting the first three PCs for use as control variables. The first three PCs explained 56.9% of the variance in covariates for the questionnaire models, and 57.6% when including smoking and caffeine for the HEP and interoceptive accuracy models. To account for multiple testing, *p*-values for the three main questionnaires and three HEP ROIs were adjusted using the BH procedure.

***1.4.4. Sensitivity Analysis: Parsimonious Interoceptive Accuracy Model***

The confirmatory interoceptive accuracy model included 11 predictors and 111 participants with non-missing interoceptive accuracy data, placing it at the lower bound of the recommended sample size per predictor (10–15 observations per predictor; Babyak, 2004). Given this ratio, there is an increased risk of overfitting, unstable estimates, and reduced power to detect smaller effects. To address this, we conducted an exploratory model comparison approach, sequentially adding predictors in order of theoretical importance. This approach allowed us to assess the incremental contribution of each predictor, evaluate model parsimony using AIC/BIC, and identify isolated effects of predictors, thereby balancing fit and complexity. The best-fitting parsimonious model was interoceptive_accuracy ~ group + sex, which captured most of the explained variance (*R²* = 0.19) while minimizing model complexity (see Table S2 for model comparison). Adding additional covariates beyond this model provided minimal gains and increased AIC/BIC, suggesting reduced efficiency.

***1.4.5. Topographical and Temporal HEP Patterns***

We performed a spatio-temporal cluster-based permutation test on HEP amplitudes to examine group differences between SSD and HC, beyond the predefined ROIs and times of interest, for the three EEG conditions separately. Data from 0.25 to 0.55 s post-R-peak were analyzed across all channels, a time window highlighted in the literature for capturing the HEP (Coll et al., 2021). Neighboring channels were defined using the adjacency matrix. Significance in this test was assessed by identifying contiguous clusters of spatio-temporal points exceeding an automatically defined threshold and evaluating their significance by comparing against a distribution of clusters obtained under the null hypothesis via 1000 random permutations. Cluster-level statistics were computed using MNE’s default method, which automatically sets cluster-forming thresholds from the test statistic distribution and controls the family-wise error rate at α = 0.05.

***1.4.6. Multivariate Brain–Body–Symptom Associations***

Within the patient sample, Partial Least Squares (PLS) regression, plsr package on R, was applied to investigate the multivariate relationship between interoceptive and physiological measures (X block) and clinical symptomatology (Y block: PANSS positive, negative, and general subscales). The X block included HEP averaged across all tasks from the CP1 and CP2 channels, which were identified in prior exploratory cluster-based permutation analyses as the most prominent channels differentiating SSD from HC, suggesting their potential clinical relevance. In addition, the X block comprised HR, HRV, interoceptive accuracy, and all questionnaire subscales, with all physiological and EEG measures averaged across tasks to capture stable, trait-like effects. To remove variance attributable to demographic and medication confounders, all interoceptive and physiological variables were residualized for age, sex, BMI, years of education, and CPZ. All variables were centered and scaled prior to analysis and two latent components were extracted. The PLS model estimates regression coefficients that describe how strongly each X variable contributes to each latent component in predicting the Y block. To assess the stability and statistical significance of these coefficients, jackknife resampling was applied, which repeatedly refits the model while leaving out one participant at a time. This procedure yields approximate t-tests for each regression coefficient, providing a robust estimate of which predictors reliably contribute to the X–Y relationship. Finally, component-level significance was assessed using 1,000 permutation tests, in which the correspondence between X and Y scores was randomly shuffled to determine whether the observed correlations between component scores exceeded what could be expected by chance. This approach allows identification of interoceptive and physiological variables that are most reliably associated with clinical symptom dimensions in SSD.

***1.4.7. Control Analyses***

To examine whether interoceptive accuracy was influenced by attentional performance, we conducted an additional linear regression analysis with interoceptive accuracy as the dependent variable and standardized attentional performance as the predictor. Attention was operationalized using the standardized composite score of the attention subtest of the BACS, with higher scores indicating better attentional performance.

To exclude the possibility that group differences in HEP were confounded by residual cardiac field artifacts (CFA) reflected in peripheral cardiac signal amplitude, we tested for group differences in mean ECG amplitude within the HEP time window (450–500 ms post–R-peak) separately for the eyes-closed, eyes-open, and HCT conditions using linear regression models with group as predictor.

2. **Supplementary Results**

For HEP analyses, task-specific EEG data-quality exclusions were applied. In the eyes-closed condition (confirmatory analysis), 1 HC dataset was excluded due to noisy EEG, yielding a final sample of 59 HC and 53 SSD participants. In the eyes-open condition, 6 HC and 3 SSD datasets were excluded, resulting in final samples of 54 HC and 50 SSD participants. In the HCT condition, 1 HC dataset was excluded, resulting in final samples of 59 HC and 53 SSD participants.

Analysis-based exclusion Criteria were as follows. Questionnaire data: Participants were excluded from questionnaire-based analyses if questionnaire data were deemed unusable during data collection or inspection. Specifically, rows were excluded when the variable excluded_questionnaires = 1, indicating that unplanned or irregular events occurred that compromised data validity (e.g., extremely rapid responding suggestive of non-engagement, failure to follow instructions, or other anomalies noted by the experimenter). Participants with missing exclusion flags were retained. This led to the exclusion of the questionnaires of a single participant from the SSD group.

Interoceptive accuracy: For analyses of interoceptive accuracy, participants were excluded if behavioral task validity criteria were not met. Specifically, individuals were excluded when either (a) excluded_hct_beh = 1, indicating irregularities during task performance (e.g., distraction, misunderstanding of instructions), or (b) HCT_counted_body = 0, indicating that participants reported counting non-bodily cues (e.g., time estimation, external signals) rather than heartbeat sensations. Only participants who counted bodily sensations and passed behavioral quality checks were included in IAcc analyses. ECG-derived parameters were merged exclusively for the HCT condition. All participants were retained for the interoceptive accuracy analysis.

EEG data: EEG data exclusions were applied at the run level, separately for eyes-closed, eyes-open, and HCT conditions.

- Run-level exclusions during preprocessing: If excluded_ec, excluded_eo, or excluded_hct was set to 1, the corresponding run was excluded prior to analysis due to data quality issues identified during preprocessing (e.g., technical failures, excessive artifacts; all participants retained).
- Epoch-level quality control: If an epoch had double R-peaks, it was excluded. Also, runs with more than 33% rejected epochs were inspected further.
  - If excessive rejection was driven by a single channel, that channel was interpolated.
  - If excessive rejection was distributed across channels, the run was excluded.
- Channel-level quality control:
  - ROI-based analyses: Runs were excluded if more than one region-of-interest (ROI) channel required interpolation. Consequently, of HC group 1 eyes-closed, 5 eyes-open, and 1 HCT run was excluded; of SSD group 3 eyes-open runs were excluded.
  - Spatio-temporal cluster analyses: Participants were excluded if more than 33% of channels were marked as bad in a given condition. Three eyes-open runs of HC were excluded.

These procedures ensured that only data of sufficient quality contributed to ROI and whole-scalp EEG analyses.

**2.1. Confirmatory Results**

***2.1.1. Self-Report and Beliefs on Bodily Perception***

To examine group differences in self-reported bodily perception, we fitted linear models and found that individuals with SSD reported a reduced conscious interoceptive body awareness as measured by MAIA total (β = -0.83, 95% CI [-1.29, -0.36], *p*_adjusted_ = 0.001), alongside increased depersonalization (CDS; β = 0.88, 95% CI [0.41, 1.35], *p*_adjusted_ = 0.001). In contrast, no group differences were observed for BPQ total scores, indicating comparable perceived bodily and autonomic symptom frequency across groups. See Table S3.

***2.1.2. Interoceptive Accuracy***

Individuals with SSD showed a non-significant trend toward lower interoceptive accuracy compared with HC (β = -0.48, 95% CI [-0.99, 0.02], *p* = 0.058, *R^2^*_adjusted_ = 0.16; see Figure 2B and Table S4).

***2.1.3. Resting-State HEP in Preregistered Right-Frontal ROIs***

No significant group differences in HEP amplitudes were observed after adjustment for covariates at any of the predefined ROI channels: Fp2 (β = 0.35, 95% CI [-0.10, 0.81], *p*_adjusted_ = 0.38), F4 (β = -0.12, 95% CI [-0.57, 0.33], *p*_adjusted_ = 0.60), or F8 (β = 0.13, 95% CI [-0.32, 0.59], *p*_adjusted_ = 0.60). Therefore, the results did not replicate the results reported by Koreki et al. (2024). See Table S5.

**2.2. Exploratory Results**

***2.2.1. Questionnaire Subscales***

Exploratory analyses of MAIA subscales revealed selective reductions in self-reported interoceptive body awareness in SSD participants. Specifically, SSD was associated with lower scores on Not Worrying (β = -0.72, 95% CI [-1.19, -0.24], *p*_adjusted_ = 0.014), Attention Regulation (β = -0.75, 95% CI [-1.21, -0.29], *p*_adjusted_ = 0.013), Emotional Awareness (β = -0.57, 95% CI [-1.05, -0.09], *p*_adjusted_ = 0.035), Self-Regulation (β = -0.67, 95% CI [-1.15, -0.19], *p*_adjusted_ = 0.014), and Trusting (β = -0.67, 95% CI [-1.14, -0.20], *p*_adjusted_ = 0.014). Other subscales, including Noticing (β = -0.22, 95% CI [-0.71, 0.27], *p*_adjusted_ = 0.38), Not Distracting (β = -0.24, 95% CI [-0.72, 0.24], *p*_adjusted_ = 0.38), Body Listening (β = -0.27, 95% CI [-0.74, 0.21], *p*_adjusted_ = 0.35), did not show significant differences between groups.

SSD was further associated with higher scores on the Supra-Diaphragmatic subscale (β = 0.64, 95% CI [0.17, 1.11], *p*_adjusted_ = 0.025), indicating increased frequency of supra-diaphragmatic bodily awareness. Subscales assessing Body Awareness (β = 0.01, 95% CI [-0.48, 0.49], *p*_adjusted_ = 0.98) and Sub-Diaphragmatic sensations (β = 0.35, 95% CI [-0.13, 0.83], *p*_adjusted_ = 0.23) did not show significant group differences. These results suggest that SSD is linked to domain-specific impairments in interoceptive awareness rather than uniform reductions across all subscales.

***2.2.2. Parsimonious Interoceptive Accuracy Model***

The model revealed a significant overall effect of group and sex. Specifically, participants in the SSD group showed lower interoceptive accuracy compared to controls (β = -0.177, 95% CI [-0.269, -0.084], *t* = -3.77, *p* = 0.0003, *R^2^*_adjusted_ = 0.18). Additionally, males showed higher interoceptive accuracy than females (β = 0.166, 95% CI [0.073, 0.258], *p* = 0.0006).

***2.2.3. Spatiotemporal HEP Patterns***

The spatio-temporal cluster permutation tests revealed no significantly different HEP clusters for the eyes-closed condition for both groups. However, for the HCT condition, a total of 14 significant spatio-temporal clusters were identified, where the SSD group showed a smaller HEP positivity amplitude. These clusters involved a time window of approximately 0.30–0.55 s post-R-peak, primarily located in the centro-parietal (CP1, CP2, P4, Pz, C3) electrodes, with additional engagement of frontal (F3, F4, FC1, FC6) and prefrontal sites (Fp1, Fp2), and occasional extensions toward occipital (O2) and temporo-parietal regions (P7, TP9). A similar pattern, though less extensive and involving fewer clusters, was observed for the eyes-open condition, where three significant spatio-temporal clusters were identified. These clusters were significant within a time window of approximately 0.37–0.53 s post-R-peak. The channels involved primarily included centro-parietal (CP1, CP2, Pz, P4, C3, P3) and frontal (F3) electrodes, with one cluster extending to a temporo-parietal site (TP10). In summary, the SSD group consistently exhibited a reduction in HEP positivity amplitude within the centro-parietal regions. Crucially, this effect was modality-dependent, being robustly observed during HCT, to a lesser extent during the eyes-open condition, and not at all when participants were in the eyes-closed condition (see Table S7 for a summary).

***2.2.4. Task Effects on HEP in the ROI***

HEP amplitude measured at Fp2 in the SSD group showed a reduction in negativity, hence a more positive value, as compared to HC (β = 0.52, 95% CI [0.14, 0.89], t = 2.71, *p*_adjusted_ = 0.02). This indicates that for this channel, there was a total effect of the group on HEP amplitudes. We did not observe any main effects or interactions for any other ROI (see Table S8, Figure S2). These analyses estimate the ROI-based total effects and interactions without covariate adjustment and are therefore reported as supplementary and exploratory.

***2.2.5. Multivariate Brain–Body–Symptom Associations***

To uncover latent relationships between interoceptive/physiological and clinical variables, we extracted two latent PLS components. Component 1 explained 7.3% of the variance in interoceptive/physiological predictors and 8.1% in PANSS scores; Component 2 explained 7.5% and 3.8%, respectively. Permutation tests indicated that neither latent component reached significance (Component 1*: p* = 0.74; Component 2: *p* = 0.74). However, several variables showed consistent, directionally meaningful jackknife estimates. PANSS negative was positively associated with BPQ supra-diaphragmatic reactivity (β = 0.10, *SE* = 0.03, *t*(9) = 3.31, *p* = 0.009) and with CDS (β = 0.12, *SE* = 0.02, *t*(9) = 5.14, *p* < 0.001), but negatively associated with MAIA Trusting (β = −0.17, *SE* = 0.06, *t(*9) = −2.95, *p* = 0.016). Jackknife tests further revealed several trend-level associations, although no predictors survived the conventional α = .05 threshold. PANSS positive symptoms showed a positive trend with interoceptive accuracy (β = 0.16, *SE* = 0.07, *t*(9) = 2.22, *p* = 0.054), CDS (β = 0.20, *SE* = 0.10, *t*(9) = 1.92, *p* = 0.086, 95% CI [−0.035, 0.433]), and MAIA body listening (β = 0.20, *SE* = 0.09, *t*(9) = 2.20, *p* = 0.055). For PANSS general psychopathology CDS again showed a trend-level positive association (β = 0.18, *SE* = 0.08, *t*(9) = 2.22, *p* = 0.054). No other predictors reached significance. See Table S9, Figure S3.

### 3.3. Control Analyses

Attentional performance significantly correlated with interoceptive accuracy, with higher attention scores associated with higher interoceptive accuracy (β = 0.09, *SE* = 0.03, *t* = 2.84, *p* = 0.007).

No significant group differences in mean ECG amplitude within the HEP time window were observed in the eyes-closed (β = 0.08, *t* = 0.41, *p* = .68), eyes open (β = 0.18, *t* = 0.88, *p* = .38), or HCT (β = 0.12, *t* = 0.63, *p* = .533) conditions. See Figure S1.

3. **Supplementary Discussion**

An additional consideration is the potential influence of antipsychotic medication on self-reported interoception. In partial correlation analyses, controlling for CPZ markedly attenuated the association between PANSS total and MAIA total scores, suggesting that medication dose may statistically explain a substantial portion of their shared variance. While this pattern does not permit causal inference, it raises the possibility that antipsychotic-related side effects (e.g., sedation, autonomic changes, or blunted affective signaling) may directly influence subjective interoceptive awareness. Alternatively, PANSS total may constitute a common underlying factor driving both higher medication dose and reduced MAIA scores, rendering CPZ an epiphenomenal outcome rather than a confounding variable. Future studies explicitly modeling medication effects, longitudinal symptom trajectories, and side-effect profiles will be required to disentangle illness-related from pharmacologically induced interoceptive alterations (See Table S10).

Attentional scores explained only a modest proportion of variance in interoceptive accuracy within the SSD group. While we could not statistically adjust for attention across groups, as it was collected only for the SSD sample, the persistence and direction of the group effect suggest that attentional deficits alone are unlikely to fully account for reduced interoceptive accuracy in SSD. Therefore, this association points more toward a shared mechanism, suggesting that there may be both domain-general (attention-related) and domain-specific (interoceptive) components underlying interoceptive accuracy. Although the HCT by Shandry et al. (1981) performance is often criticized for being confounded by prior knowledge and estimation strategies, we minimized these influences by providing strict instructions, limiting estimation cues, and statistically verifying that explicit knowledge of HR did not account for performance. The reduced accuracy in SSD has also been reported using interoceptive tasks less prone to cognitive estimation (e.g., heartbeat discrimination, respiratory detection), further supporting the notion that the deficit extends beyond attentional confounds and may reflect a genuine alteration in interoceptive signal processing.

Although the confirmatory analyses testing Hypothesis 1.3 did not survive covariate adjustment and correction for multiple comparisons, exploratory analyses revealed condition-independent alterations in HEP amplitude. Specifically, among the hypothesis-driven channels (Fp2, F4, and F8; based on Koreki et al., 2024), only Fp2 showed a significant reduction in HEP negativity in SSD across all three EEG conditions in the exploratory parsimonious model, with a medium-to-large effect size (see Section 3.2.3). This indicates a total diagnostic group effect on HEP that generalizes across task contexts, but is likely too subtle in the eyes-closed condition to remain significant under strict confirmatory adjustment.

The absence of this effect in the full confirmatory model likely reflects differences in statistical control, including the inclusion of multiple covariates and correction for multiple comparisons, as well as partial collinearity between diagnostic group and several covariates, which may have reduced estimation stability and statistical power. When examining bivariate associations, caffeine intake showed a modest association with HEP amplitude, whereas BMI and cardiac-related variables did not demonstrate consistent relationships. This pattern argues against residual cardiac field artifacts as a primary explanation and instead suggests that non-cardiac physiological or state-related factors may exert subtle modulatory influences on cortical HEP generation. Notably, caffeine intake did not differ between groups (Table 1), indicating a general physiological modulation rather than a group-specific confound. Importantly, control analyses revealed no group differences in ECG amplitude within the confirmatory post–R-peak time window, supporting the interpretation that observed HEP differences reflect genuine neural activity rather than residual cardiac contamination.

Additional evidence for reduced HEP amplitude in SSD emerged from the exploratory cluster-based permutation analyses (Section 3.2.4). During the HCT, SSD participants exhibited markedly reduced HEP positivity, reflected by 14 significant spatiotemporal clusters over centro-parietal regions between 0.40–0.55 s post–R-peak. A similar but weaker pattern was observed during the eyes-open condition, with three significant clusters within the same time window, indicating convergent but less robust evidence for attenuated HEP responses in SSD. The fact that the most robust group differences emerged during HCT, followed by eyes-open resting state, but were not present during eyes-closed resting state, can be interpreted through several mechanisms.

First, both HCT and eyes-open fixation impose greater attentional and exteroceptive demands, while eyes-closed rest involves minimal executive or visual processing. Visual processing alterations are well-documented in SSD, including retinal dysfunction (Boudriot et al., 2024; Friedel et al., 2022; Kazakos & Karageorgiou, 2020; Silverstein et al., 2020), oculomotor control (Levy et al., 2010; O’Driscoll & Callahan, 2008), and alterations observable even in high-risk populations (Obyedkov et al., 2019). Such disruptions may interfere with the integration of interoceptive and exteroceptive information during attentionally or visually demanding states, amplifying HEP group differences. In contrast, eyes-closed rest eliminates exteroceptive visual input and minimizes saccadic scanning (Martinez-Conde et al., 2004), which may help explain the absence of significant HEP differences in this condition.

Second, eyes-closed rest is characterized by unconstrained spontaneous thought processes, where attention freely fluctuates between bodily sensations, stimulus-independent thoughts, and mind-wandering. SSD is associated with atypical default mode network (DMN) dynamics and increased spontaneous self-generated cognitive activity (Alderson-Day et al., 2016; Northoff & Duncan, 2016; Wang et al., 2024), including intrusive inner speech (Alderson-Day et al., 2016), excessive mind-wandering (Chen et al., 2018), and hyper-reflexive self-focus (Sass & Parnas, 2003). Such tendencies may induce considerable variability in focus and draw attention away from bodily sensations during eyes-closed rest, reducing the sensitivity of HEP to detect group-level interoceptive differences.

Finally, the HCT imposes the strongest interoceptive focus, requiring sustained attention to heartbeat sensations with minimal exteroceptive competition or saccadic interference. This creates a highly constrained interoceptive context in which heartbeat signals are most salient, allowing HEP alterations to emerge most clearly. Although the tasks were administered in a fixed sequence, and modest sequence-related influences, e.g., increasing familiarity or reduced anxiety over time, cannot be entirely ruled out, the progression of group differences closely and consistently followed the increasing interoceptive and attentional demands of the tasks. This convergence makes it unlikely that sequence or familiarity effects alone account for the observed pattern.

These findings emphasize that interoception is not a static or context-independent construct, but a dynamically modulated process that varies according to attentional demands, task context, and exteroceptive input (Ainley et al., 2016; Seth & Friston, 2016). The absence of significant HEP differences in eyes-closed condition suggests that a passive, unconstrained resting condition may not be sufficiently sensitive to capture interoceptive alterations in SSD. By contrast, both HCT and eyes-open rest provide stronger contextual scaffolding, through directed bodily attention or sustained visual engagement and saccadic activity, which appears to unmask disrupted interoceptive processing in SSD. Thus, interoceptive differences in SSD may emerge most clearly when the interaction between interoceptive and exteroceptive systems is challenged, rather than under purely passive conditions.

The direction of these effects are consistent with previous work reporting attenuated HEP amplitudes in SSD (Koreki et al., 2024). While the authors interpreted their findings as increased HEP in SSD, their plotted waveforms suggest that the effect was characterized by a reduction in negativity, resulting in amplitudes that were closer to zero. Importantly, both their data and ours demonstrated less spatially distinct HEP topographies in SSD, which may reflect reduced neural differentiation and weakened representation of interoceptive signals.

Unlike Koreki et al., who observed attenuated HEP responses in SSD during passive eyes-closed rest, our results show that interoceptive alterations in SSD become more pronounced under increased precision demands (HCT) and when interoceptive–exteroceptive integration is required (eyes-open). This supports predictive coding accounts (Yao & Thakkar, 2022b) suggesting that interoceptive disruption in SSD reflects context-dependent alterations in precision weighting, rather than a global loss of interoceptive sensitivity. Specifically, reduced cortical expression of HEP may reflect decreased precision weighting of visceral signals, i.e., afferent cardiac inputs are assigned insufficient confidence, leading to weaker cortical updating of bodily states, consistent with a broader account of aberrant precision signaling in SSD (Barrett & Simmons, 2015; Seth & Friston, 2016; Sterzer et al., 2018).

**Supplementary Tables and Figures**

Table S1

*Participant Characteristics*

| **Variable** | **SSD (%)** | **HC (%)** | ***χ²*** | ***p*** |
| --- | --- | --- | --- | --- |
| *Questionnaires & Interoceptive Accuracy* | | | | |
| BPQ Total | 95.47 ± 24.04 | 89.57 ± 26.85 | -1.23 | 0.22 |
| MAIA_total | 21.35 ± 5.26 | 24.89 ± 4.89 | 3.69 | **< 0.001** |
| CDS | 30.68 ± 31.32 | 13.78 ± 19.70 | -3.38 | **0.0001** |
| BPQ Body Awareness | 63.83 ± 20.58 | 62.75 ± 24.15 | -0.26 | 0.80 |
| BPQ Autonomic | 31.40 ± 8.55 | 26.68 ± 7.00 | -3.18 | **0.002** |
| BPQ Subdiaphragmatic | 9.77 ± 3.30 | 8.80 ± 2.75 | -1.69 | 0.09 |
| BPQ Supradiaphragmatic | 23.00 ± 6.86 | 19.25 ± 5.26 | -3.23 | **0.002** |
| MAIA Noticing | 3.00 ± 1.10 | 3.27 ± 1.09 | 1.31 | 0.19 |
| MAIA Not Distracting | 2.27 ± 1.00 | 2.65 ± 0.99 | 2.05 | **0.04** |
| MAIA Not Worrying | 2.52 ± 1.03 | 3.00 ± 0.97 | 2.56 | **0.02** |
| MAIA Attention Regulation | 2.55 ± 1.07 | 3.25 ± 0.93 | 3.67 | **< 0.001** |
| MAIA Emotional Awareness | 3.05 ± 1.13 | 3.42 ± 1.00 | 1.84 | 0.07 |
| MAIA Self Regulation | 2.49 ± 1.24 | 2.96 ± 0.94 | 2.27 | **0.03** |
| MAIA Body Listening | 2.13 ± 1.19 | 2.36 ± 0.99 | 1.14 | 0.26 |
| MAIA Trusting | 3.35 ± 1.14 | 4.04 ± 0.84 | 3.6 | **< 0.001** |
| Interoceptive Accuracy | 0.28 ± 0.21 | 0.45 ± 0.29 | 3.47 | **< 0.001** |
| *Cardiac Variables (Eyes-Closed)* | | | | |
| Heart Rate (BPM) | 79.91 ± 12.68 | 71.76 ± 11.20 | -3.52 | **< 0.001** |
| Heart Rate Variability (RMSSD, ms) | 7574.18 ± 7318.63 | 11455.66 ± 9352.18 | 2.43 | **0.02** |
| R-peak Amplitude (mV) | 0.94 ± 0.43 | 1.04 ± 0.61 | 0.92 | 0.36 |
| QT Interval (ms) | 331.64 ± 53.16 | 362.45 ± 44.86 | 3.24 | **0.002** |
| QTc Interval (ms) | 378.36 ± 48.31 | 392.18 ± 29.56 | 1.76 | 0.08 |
| *HEP* | | | | |
| Fp2 Mean Amplitude (μV) | 0.11 ± 2.48 | -0.61 ± 0.94 | -1.92 | 0.06 |
| F4 Mean Amplitude (μV) | -0.14 ± 0.83 | -0.26 ± 0.87 | -0.74 | 0.46 |
| F8 Mean Amplitude (μV) | -0.31 ± 1.10 | -0.59 ± 1.74 | -1 | 0.32 |

Table S2

*Interoceptive Accuracy Model Comparison*

| **Model** | **R_squared** | **Adjusted**  **R_squared** | **AIC** | **BIC** | **R_squared**  **Change** | **Adjusted**  **R_squared**  **Change** |
| --- | --- | --- | --- | --- | --- | --- |
| intercept_only | 0 | 0 | 28.38 | 33.80 | NA | NA |
| interoceptive_accuracy ~ 1 + group | 0.10 | 0.09 | 18.64 | 26.77 | 0.10 | 0.09 |
| interoceptive_accuracy ~ 1 + group + sex | 0.19 | 0.18 | 8.41 | 19.25 | 0.09 | 0.09 |
| interoceptive_accuracy ~ 1 + group + sex + education_years | 0.21 | 0.20 | 7.13 | 20.68 | 0.02 | 0.02 |
| interoceptive_accuracy ~ 1 + group + sex + education_years + MAIA_not_worrying | 0.23 | 0.20 | 7.67 | 23.92 | 0.01 | 0.003 |
| interoceptive_accuracy ~ 1 + group + sex + education_years + MAIA_not_worrying + heart_rate_bpm | 0.22 | 0.18 | 11.31 | 30.02 | -0.01 | -0.02 |
| interoceptive_accuracy ~ 1 + group + sex + education_years + MAIA_not_worrying + heart_rate_bpm + smoker | 0.22 | 0.18 | 12.97 | 34.35 | 0.002 | -0.01 |
| interoceptive_accuracy ~ 1 + group + sex + education_years + MAIA_not_worrying + heart_rate_bpm + smoker + age | 0.23 | 0.17 | 14.23 | 38.28 | 0.01 | -0.003 |
| interoceptive_accuracy ~ 1 + group + sex + education_years + MAIA_not_worrying + heart_rate_bpm + smoker + age + body_mass_index | 0.23 | 0.17 | 15.57 | 42.30 | 0.01 | -0.003 |
| interoceptive_accuracy ~ 1 + group + sex + education_years + MAIA_not_worrying + heart_rate_bpm + smoker + age + body_mass_index + had_caffeine | 0.23 | 0.16 | 17.34 | 46.74 | 0.001 | -0.007 |
| interoceptive_accuracy ~ 1 + group + sex + education_years + MAIA_not_worrying + heart_rate_bpm + smoker + age + body_mass_index + had_caffeine + knows_heartrate | 0.23 | 0.15 | 19.23 | 51.31 | 0.0007 | -0.008 |

Table S3

*Confirmatory Self-report Analysis: Linear Model Results*

| **outcome** | **predictor** | **estimate** | **std_error** | **t_value** | **p_value** | **CI_lower** | **CI_upper** | **eta_sq** | **partial_eta_sq** | **p_adjusted** |
| --- | --- | --- | --- | --- | --- | --- | --- | --- | --- | --- |
| BPQ_total | (Intercept) | 116.113 | 17.792 | 6.526 | 0 | 80.838 | 151.388 | NA | NA | 0.388 |
| BPQ_total | groupSSD | 5.396 | 6.225 | 0.867 | 0.388 | -6.945 | 17.737 | NA | NA | NA |
| BPQ_total | age | -0.236 | 0.21 | -1.126 | 0.263 | -0.652 | 0.18 | 0.011 | 0.012 | NA |
| BPQ_total | Education_  years | -1.022 | 0.663 | -1.541 | 0.126 | -2.336 | 0.293 | 0.021 | 0.022 | NA |
| BPQ_total | body_mass_index | 0.132 | 0.671 | 0.196 | 0.845 | -1.198 | 1.461 | 0 | 0 | NA |
| BPQ_total | sexm | -5.729 | 4.953 | -1.157 | 0.25 | -15.549 | 4.091 | NA | NA | NA |
| MAIA_total | (Intercept) | 27.843 | 3.561 | 7.818 | 0 | 20.783 | 34.904 | NA | NA | NA |
| MAIA_total | groupSSD | -4.204 | 1.246 | -3.374 | 0.001 | -6.675 | -1.734 | NA | NA | 0.002 |
| MAIA_total | age | 0.031 | 0.042 | 0.742 | 0.46 | -0.052 | 0.114 | 0.004 | 0.005 | NA |
| MAIA_total | Education_  years | -0.252 | 0.133 | -1.9 | 0.06 | -0.515 | 0.011 | 0.029 | 0.033 | NA |
| MAIA_total | body_mass_index | 0.035 | 0.134 | 0.259 | 0.796 | -0.231 | 0.301 | 0.001 | 0.001 | NA |
| MAIA_total | sexm | -0.355 | 0.991 | -0.358 | 0.721 | -2.32 | 1.611 | NA | NA | NA |
| CDS | (Intercept) | 46.084 | 16.098 | 2.863 | 0.005 | 14.168 | 78.001 | NA | NA | NA |
| CDS | groupSSD | 20.076 | 5.632 | 3.565 | 0.001 | 8.91 | 31.243 | NA | NA | 0.002 |
| CDS | age | -0.363 | 0.19 | -1.915 | 0.058 | -0.739 | 0.013 | 0.029 | 0.033 | NA |
| CDS | Education_  years | -0.331 | 0.6 | -0.551 | 0.583 | -1.52 | 0.859 | 0.002 | 0.003 | NA |
| CDS | body_mass_index | -0.662 | 0.607 | -1.091 | 0.278 | -1.865 | 0.541 | 0.009 | 0.011 | NA |
| CDS | sexm | 2.885 | 4.481 | 0.644 | 0.521 | -6 | 11.77 | NA | NA | NA |

Table S4

*Confirmatory Interoceptive Accuracy Analysis: Linear Model Results*

| **predictor** | **outcome** | **std_estimate** | **std_95CI_low** | **std_95CI_high** | **t_value** | **p_value** |
| --- | --- | --- | --- | --- | --- | --- |
| (Intercept) | interoceptive_accuracy | -0.12 | -0.51 | 0.27 | 0.74 | 0.46 |
| groupSSD | interoceptive_accuracy | -0.48 | -0.99 | 0.02 | -1.92 | 0.06 |
| age | interoceptive_accuracy | -0.08 | -0.29 | 0.13 | -0.75 | 0.45 |
| sexm | interoceptive_accuracy | 0.55 | 0.16 | 0.94 | 2.81 | 0.01 |
| education_years | interoceptive_accuracy | 0.19 | -0.01 | 0.38 | 1.88 | 0.06 |
| body_mass_index | interoceptive_accuracy | -0.09 | -0.35 | 0.17 | -0.7 | 0.49 |
| heart_rate_bpm | interoceptive_accuracy | 0.01 | -0.2 | 0.21 | 0.06 | 0.95 |
| smoker1 | interoceptive_accuracy | 0.11 | -0.34 | 0.56 | 0.48 | 0.63 |
| had_caffeine1 | interoceptive_accuracy | 0.01 | -0.41 | 0.42 | 0.03 | 0.98 |
| knows_heartrate1 | interoceptive_accuracy | 0.07 | -0.34 | 0.48 | 0.35 | 0.73 |
| MAIA_not_worrying | interoceptive_accuracy | 0.1 | -0.1 | 0.29 | 0.97 | 0.34 |

Table S5

*Confirmatory HEP Analysis: Linear Model Results*

| **predictor** | **outcome** | **std_estimate** | **std_95CI_low** | **std_95CI_high** | **t_value** | **p_value** | **p_adjusted** |
| --- | --- | --- | --- | --- | --- | --- | --- |
| (Intercept) | Fp2_mean_amplitude | -0.16 | -0.44 | 0.12 | -2.2 | 0.03 |  |
| groupSSD | Fp2_mean_amplitude | 0.35 | -0.1 | 0.81 | 1.54 | 0.13 | 0.38 |
| PC1 | Fp2_mean_amplitude | -0.02 | -0.24 | 0.2 | -0.16 | 0.88 |  |
| PC2 | Fp2_mean_amplitude | 0.01 | -0.18 | 0.21 | 0.14 | 0.89 |  |
| PC3 | Fp2_mean_amplitude | -0.06 | -0.25 | 0.13 | -0.65 | 0.51 |  |
| (Intercept) | F4_mean_amplitude | 0.06 | -0.22 | 0.33 | -1.3 | 0.2 |  |
| groupSSD | F4_mean_amplitude | -0.12 | -0.57 | 0.33 | -0.53 | 0.6 | 0.6 |
| PC1 | F4_mean_amplitude | -0.17 | -0.39 | 0.05 | -1.57 | 0.12 |  |
| PC2 | F4_mean_amplitude | 0.13 | -0.06 | 0.33 | 1.34 | 0.18 |  |
| PC3 | F4_mean_amplitude | -0.18 | -0.37 | 0.01 | -1.88 | 0.06 |  |
| (Intercept) | F8_mean_amplitude | -0.06 | -0.35 | 0.22 | -2.6 | 0.01 |  |
| groupSSD | F8_mean_amplitude | 0.13 | -0.32 | 0.59 | 0.58 | 0.56 | 0.6 |
| PC1 | F8_mean_amplitude | -0.08 | -0.3 | 0.14 | -0.7 | 0.49 |  |
| PC2 | F8_mean_amplitude | -0.06 | -0.26 | 0.14 | -0.57 | 0.57 |  |
| PC3 | F8_mean_amplitude | -0.04 | -0.23 | 0.16 | -0.38 | 0.71 |  |

Table S6

*Interoceptive Associations with PANSS Total*

| **x** | **y** | **estimate** | **p_value** | **t_statistic** | **df** | **CI_lower** | **CI_upper** | **p_adj_BH** |
| --- | --- | --- | --- | --- | --- | --- | --- | --- |
| **Questionnaire-based Mesures** | | | | | | | | |
| panss_total | MAIA_total | -0.23 | 0.12 | -1.6 | 3 | -0.48 | 0.06 | 0.12 |
| panss_total | BPQ_total | 0.25 | 0.09 | 1.75 | 3 | -0.04 | 0.49 | 0.12 |
| panss_total | CDS | 0.55 | <0.001 | 4.55 | 3 | 0.32 | 0.72 | <0.001 |
| **Behavioral Interoceptive Accuracy Measure** | | | | | | | | |
| panss_total | interoceptive_accuracy | 0.11 | 0.47 | 0.72 | 3 | -0.2 | 0.41 | - |
| **EEG HEP ROI Amplitudes** | | | | | | | | |
| panss_total | Fp2_mean_amplitude | -0.06 | 0.67 | -0.43 | 3 | -0.34 | 0.23 | 0.94 |
| panss_total | F4_mean_amplitude | -0.13 | 0.37 | -0.91 | 3 | -0.41 | 0.16 | 0.94 |
| panss_total | F8_mean_amplitude | 0.01 | 0.94 | 0.08 | 3 | -0.28 | 0.3 | 0.94 |

Table S7

*Spatio-temporal Cluster-based Permutation Test Results: Clusters Comparing HEP for SSD and HC*

| **Cluster ID** | **Channels Involved** | **Time Range (s)** | ***p*-value** |
| --- | --- | --- | --- |
| *Heartbeat Counting Task Resting State HEP* | | | |
| **20** | O2, CP1, CP2, FC1, P7, C3, Cz, Pz, P4, FC6, Fp1, Fp2 | 0.306 - 0.542 | 0.018 |
| **24** | O2, CP1, CP2, FC1, P7, C3, Cz, Pz, P4, FC6, F4, Fp1, Fp2 | 0.310 - 0.546 | 0.018 |
| **48** | CP1, CP2, Fp2, FC1, P7, Pz, P4, FC6, Fp1 | 0.338 - 0.534 | 0.023 |
| **52** | CP1, CP2, Fp2, FC1, P7, Pz, P4, FC6, Fp1 | 0.342 - 0.538 | 0.027 |
| **57** | Cz, TP9, CP1, CP2, Fp2, F3, FC6, C3, P3, Pz, P4 | 0.350 - 0.542 | 0.03 |
| **71** | TP9, CP1, CP2, Fp2, F3, Pz, P4 | 0.366 - 0.510 | 0.044 |
| **74** | TP9, CP1, CP2, Fp2, F3, Pz, P4 | 0.370 - 0.514 | 0.042 |
| **78** | TP9, CP1, CP2, Fp2, F3, Pz, P4 | 0.374 - 0.518 | 0.036 |
| **81** | TP9, FC1, CP1, CP2, Fp2, C3, Cz, Pz, P4, F3, O2, FC6, P3, F4 | 0.378 - 0.542 | 0.01 |
| **84** | TP9, CP1, CP2, Fp2, C3, P4, F3, FC6, P3, Pz, F4 | 0.382 - 0.546 | 0.012 |
| **88** | TP9, CP1, CP2, Fp2, C3, F3, FC6, P3, Pz, P4, F4 | 0.386 - 0.542 | 0.02 |
| **92** | TP9, CP1, CP2, Fp2, C3, F3, FC6, P3, Pz, P4, F4 | 0.390 - 0.546 | 0.018 |
| **95** | TP9, CP1, CP2, Fp2, F3, FC6, C3, P3, Pz, P4 | 0.394 - 0.538 | 0.03 |
| **116** | CP1, CP2, Fp2, F3, FC6, C3, P3, Pz, P4 | 0.450 - 0.546 | 0.042 |
| *Eyes-Open Resting State HEP* | | | |
| **54** | F3, Pz, P4, CP1, CP2, C3, P3 | 0.366 - 0.518 | 0.038 |
| **57** | F3, Pz, P4, CP1, CP2, C3, P3 | 0.370 - 0.522 | 0.041 |
| **60** | F3, Pz, P4, CP1, CP2, TP10, C3, P3 | 0.374 - 0.526 | 0.04 |

Figure S1

*Control Analysis: ECG Waveform during Eyes-Closed Rest Showing No Group Differences*

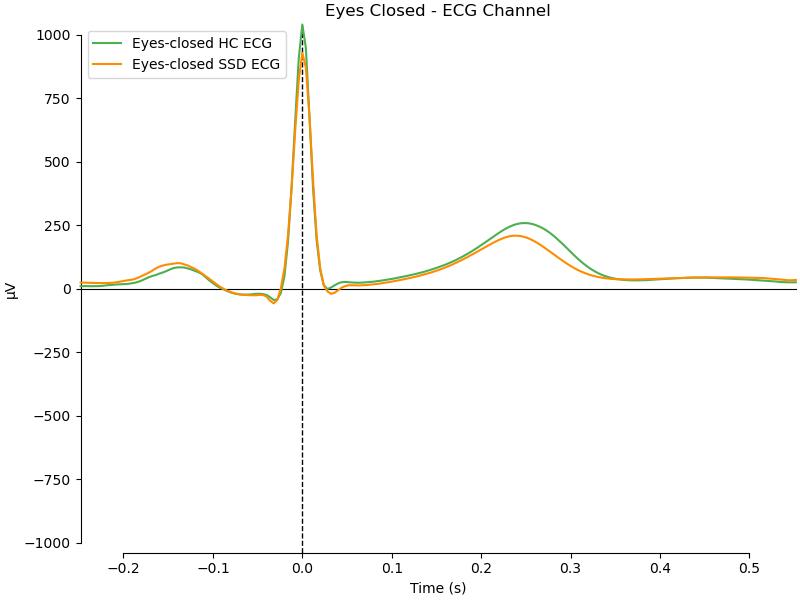

Table S8

*Group-Task HEP Interactions in ROI: Linear Model Results*

| **predictor** | **outcome** | **std_estimate** | **std_95CI_low** | **std_95CI_high** | **t** | **p** | **Adjusted**  **p_value** |
| --- | --- | --- | --- | --- | --- | --- | --- |
| (Intercept) | Fp2_mean_amplitude | -0.27 | -0.55 | 0 | -2.33 | 0.02 |  |
| groupSSD | Fp2_mean_amplitude | 0.52 | 0.14 | 0.89 | 2.71 | 0.01 | 0.02 |
| taskeyes-open | Fp2_mean_amplitude | 0.24 | -0.13 | 0.61 | 1.29 | 0.2 |  |
| taskhct | Fp2_mean_amplitude | -0.06 | -0.42 | 0.3 | -0.35 | 0.73 |  |
| sexm | Fp2_mean_amplitude | 0.09 | -0.13 | 0.31 | 0.78 | 0.43 |  |
| age | Fp2_mean_amplitude | 0 | -0.12 | 0.11 | -0.08 | 0.94 |  |
| groupSSD:  taskeyes-open | Fp2_mean_amplitude | -0.3 | -0.83 | 0.24 | -1.08 | 0.28 |  |
| groupSSD:  taskhct | Fp2_mean_amplitude | -0.14 | -0.67 | 0.39 | -0.51 | 0.61 |  |
| (Intercept) | F4_mean_amplitude | 0 | -0.28 | 0.28 | -1.28 | 0.2 |  |
| groupSSD | F4_mean_amplitude | 0.12 | -0.26 | 0.5 | 0.61 | 0.54 | 0.54 |
| taskeyes-open | F4_mean_amplitude | 0.09 | -0.29 | 0.46 | 0.46 | 0.65 |  |
| taskhct | F4_mean_amplitude | -0.12 | -0.48 | 0.25 | -0.64 | 0.52 |  |
| sexm | F4_mean_amplitude | -0.1 | -0.32 | 0.13 | -0.85 | 0.39 |  |
| age | F4_mean_amplitude | 0.02 | -0.1 | 0.13 | 0.31 | 0.76 |  |
| groupSSD:  taskeyes-open | F4_mean_amplitude | -0.06 | -0.61 | 0.49 | -0.22 | 0.83 |  |
| groupSSD:  taskhct | F4_mean_amplitude | 0.12 | -0.42 | 0.66 | 0.42 | 0.67 |  |
| (Intercept) | F8_mean_amplitude | -0.19 | -0.47 | 0.09 | -1.28 | 0.2 |  |
| groupSSD | F8_mean_amplitude | 0.24 | -0.14 | 0.62 | 1.22 | 0.22 | 0.33 |
| taskeyes-open | F8_mean_amplitude | 0.19 | -0.18 | 0.56 | 1.01 | 0.31 |  |
| taskhct | F8_mean_amplitude | 0.11 | -0.26 | 0.48 | 0.59 | 0.55 |  |
| sexm | F8_mean_amplitude | 0.02 | -0.2 | 0.24 | 0.16 | 0.87 |  |
| age | F8_mean_amplitude | -0.07 | -0.19 | 0.04 | -1.28 | 0.2 |  |
| groupSSD:  taskeyes-open | F8_mean_amplitude | -0.12 | -0.67 | 0.43 | -0.43 | 0.66 |  |
| groupSSD:  taskhct | F8_mean_amplitude | -0.07 | -0.61 | 0.47 | -0.24 | 0.81 |  |

Figure S2

*Group-Task ROI HEP Plots*

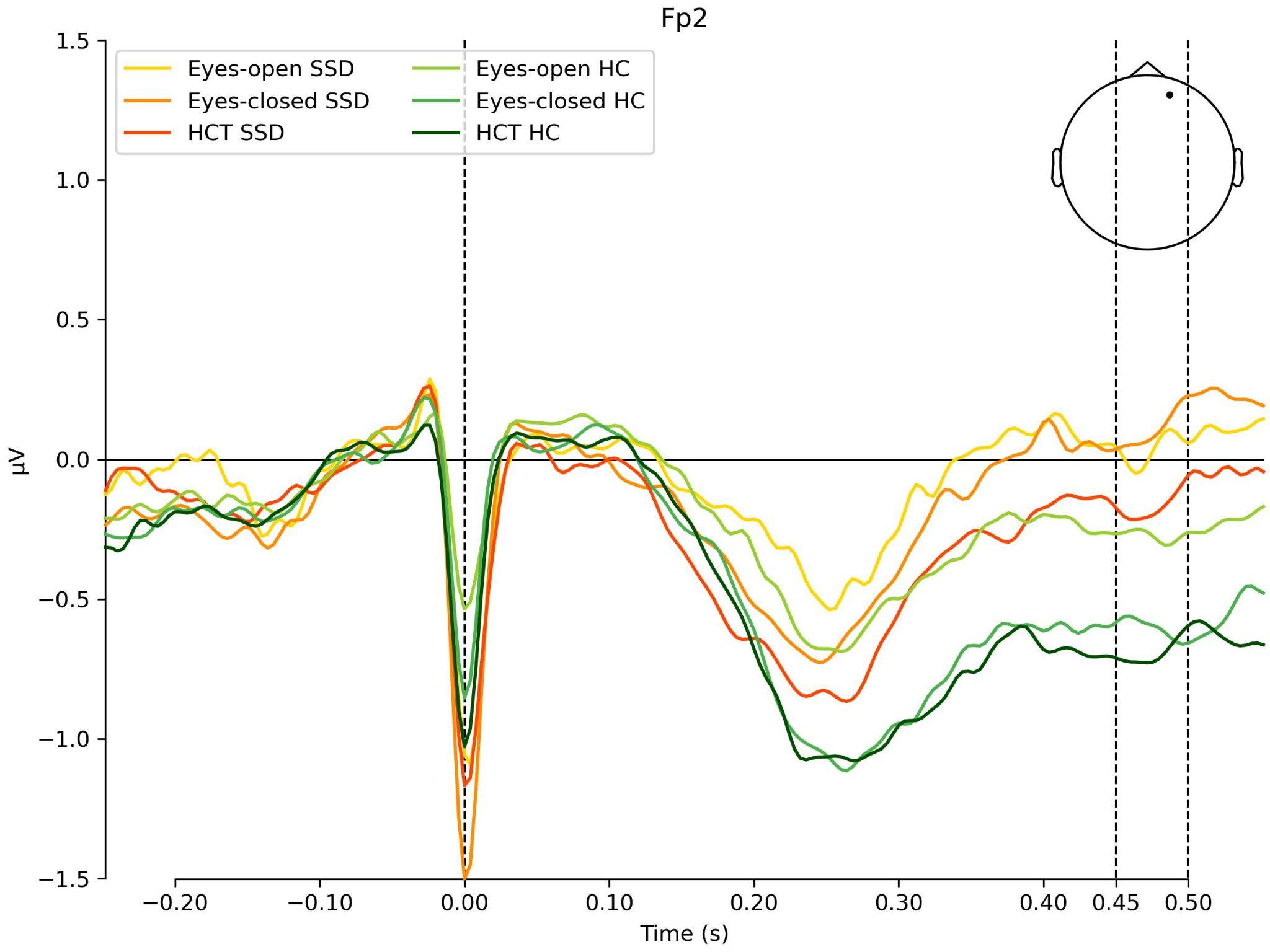

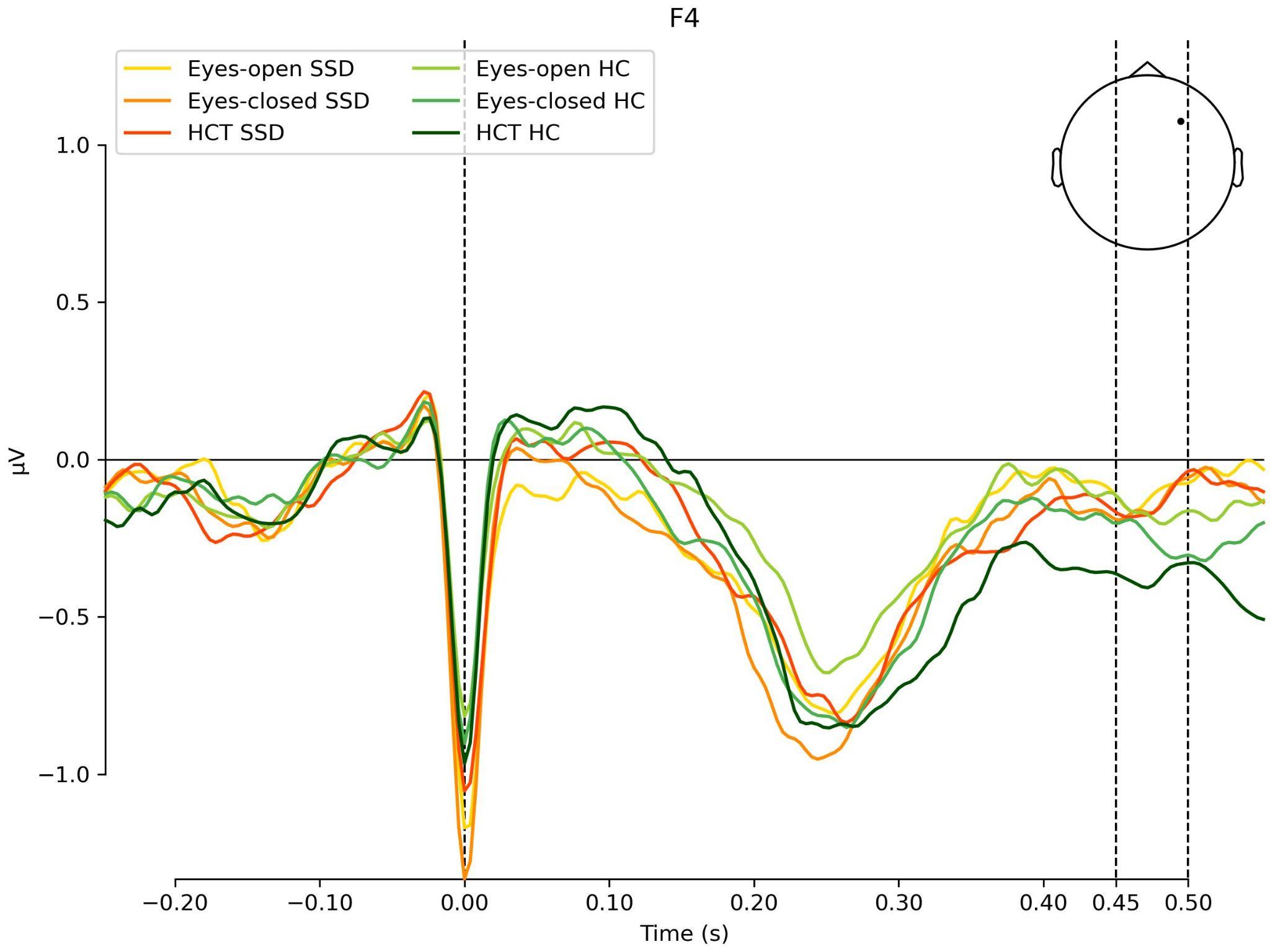

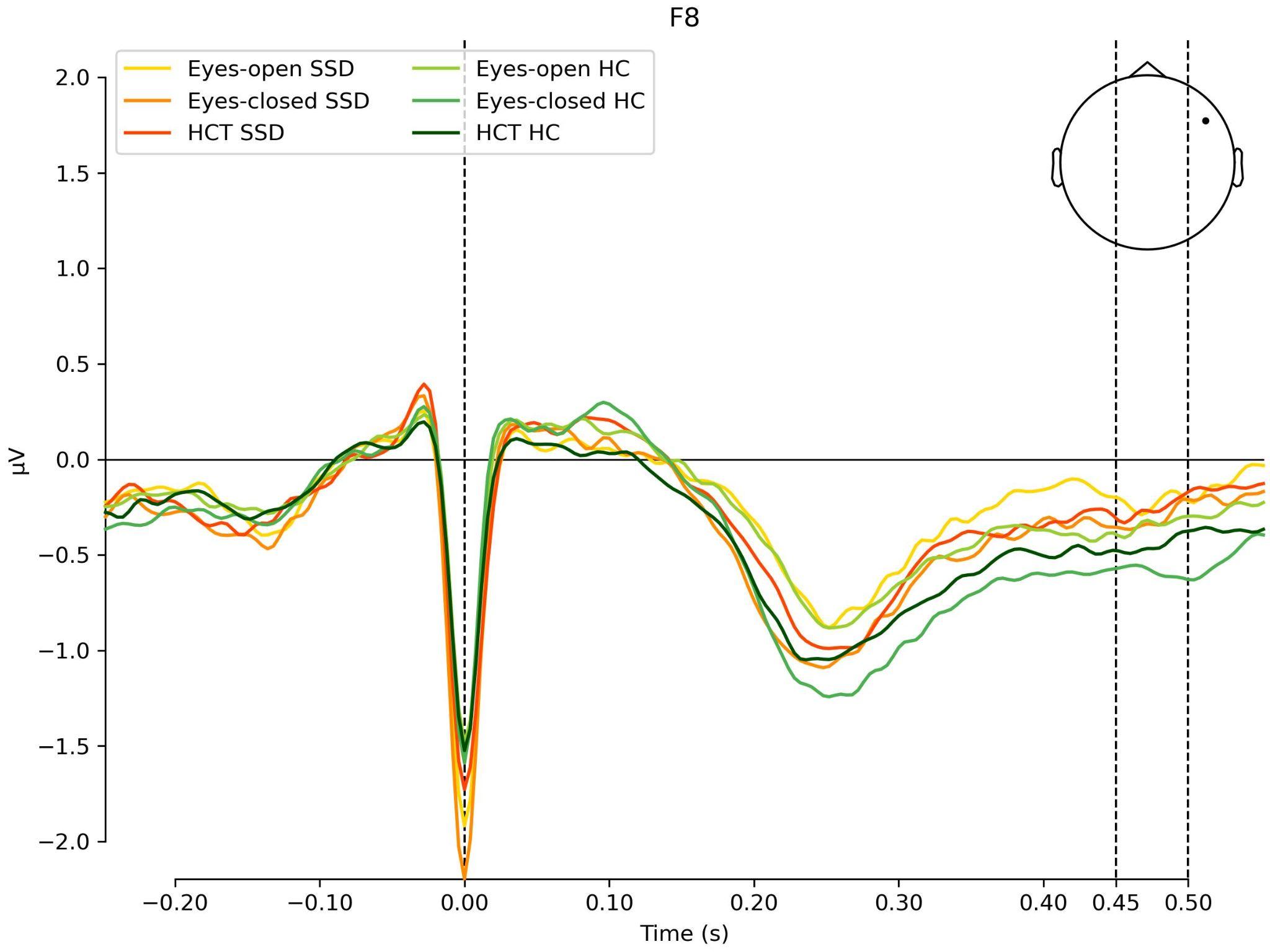

Table S9

*PLS Regression Results*

A. *Y-block Loadings (Clinical Variables): PLS loadings for PANSS dimensions*

| **Clinical variable (Y-block)** | **Comp 1** | **Comp 2** |
| --- | --- | --- |
| PANSS positive | −0.27 | 0.26 |
| PANSS negative | −0.30 | −0.18 |
| PANSS general | −0.29 | 0.11 |

B. *Jackknife Associations Between Interoceptive Predictors and PANSS Dimensions**

**PANSS Negative**

| **Predictor** | **β** | **SE** | **t** | **p** |
| --- | --- | --- | --- | --- |
| BPQ supra-diaphragmatic reactivity | 0.10 | 0.03 | 3.31 | **0.009** |
| CDS (depersonalization) | 0.12 | 0.02 | 5.14 | **<0.001** |
| MAIA trusting | −0.17 | 0.06 | −2.95 | **0.016** |

**PANSS Positive**

| **Predictor** | **β** | **SE** | **t** | **p** |
| --- | --- | --- | --- | --- |
| Interoceptive accuracy | 0.16 | 0.07 | 2.22 | 0.054 |
| CDS | 0.20 | 0.10 | 1.92 | 0.086 |
| MAIA body listening | 0.20 | 0.09 | 2.20 | 0.055 |

**PANSS General**

| **Predictor** | **β** | **SE** | **t** | **p** |
| --- | --- | --- | --- | --- |
| CDS | 0.18 | 0.08 | 2.22 | 0.054 |

*Only directionally interpretable jackknife estimates are shown.

Figure S3

*PLS Regression Results: Visualization of Loadings Relating Symptom Domains to Interoceptive Domains*

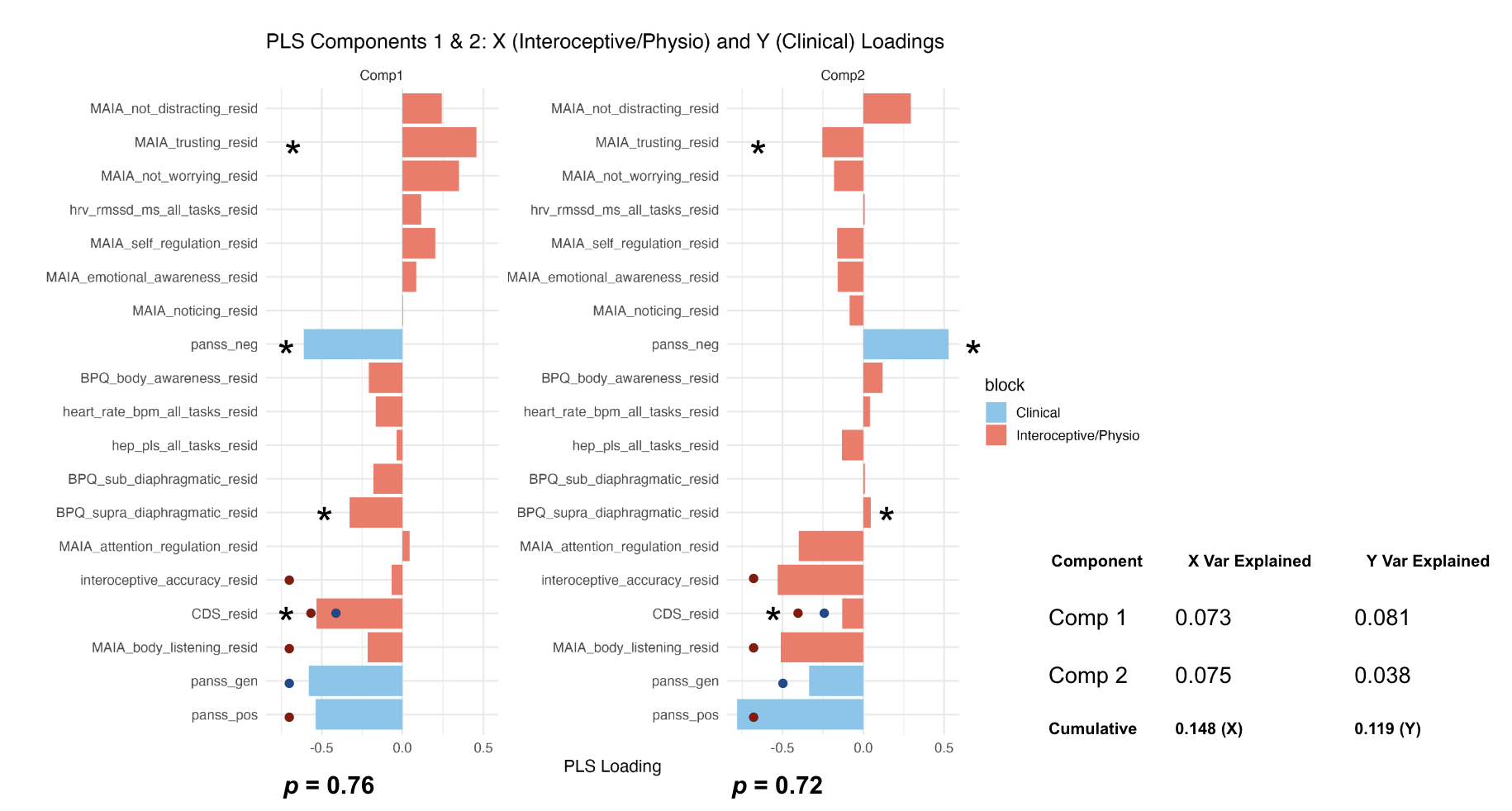

Table S10

*Relationship Between PANSS Total, MAIA Total, and CPZ*

| **Model** | **Predictor → Outcome** | **β (Estimate)** | **p-value** | **R²** |
| --- | --- | --- | --- | --- |
| 0 | PANSS → MAIA (unadjusted) | −0.13 | .031 | .09 |
| 1 | PANSS → MAIA (covariates) | −0.12 | .048 | .14 |
| 2 | PANSS → MAIA (covariates+ CPZ) | −0.07 | .232 | .30 |
| 3 | CPZ → MAIA | −0.0069 | < .001 | .21 |
| 4 | CPZ → PANSS | +0.0107 | .035 | .09 |

*Notes.* All models estimated using linear regression. Covariates included age, sex, education years, and BMI where indicated. CPZ = chlorpromazine equivalents.

Figure S4

*HEP Topography in the Confirmatory Time Window for Eyes-Closed Resting State*

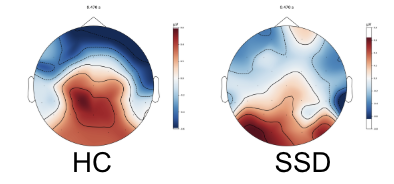
